# Genes/variants for diagnostic testing and pre-clinical research in autism spectrum disorder

**DOI:** 10.64898/2026.06.19.26356063

**Authors:** Nelson Bautista Salazar, Olivia Rennie, Worrawat Engchuan, Julian Moran, Vinicius Furlan, Xiaopu Zhou, Natalia Rivera-Alfaro, Jennifer L. Howe, Ny Hoang, Kristian Torres-Bonilla, Kristof Bosovicar, Katlin Brauer Massirer, Carl Laflamme, Aled Edwards, Karun K. Singh, Sangyoon Y. Ko, Marla Mendes de Aquino, Jacob A.S. Vorstman, Stephen W. Scherer

## Abstract

**Purpose:** Genetic discoveries have provided etiological insight into autism spectrum disorder (ASD) such that genetic testing has become standard of care. To date, consensus about which genes are robustly associated with autism liability, and which are not, is inconsistent. Consequently, different curated ASD gene lists are applied in diagnostic testing, pre-clinical model development, and the design of precision therapeutics.

**Methods:** We address this issue using the Evaluation of Autism Gene Link Evidence (EAGLE) framework, which allows for a protocolized and replicable curation of genes. Additionally, we compared the functional-and expression-characteristics of EAGLE-curated genes.

**Results:** We curated 222 genes and found 78 genes with definitive EAGLE-evidence for association with autism: 43 with moderate evidence, and 99 with limited evidence for a role in ASD (noting all 222 genes are associated with the broader category of neurodevelopmental disorders (NDDs)). The top 10 EAGLE-scoring genes are *NRXN1*-*SCN2A*-*MECP2*-*CHD8*-*RNU4-2*-*DDX3X*-*SHANK3*-*PTEN*-*FOXP1,* and *MBD5*.

**Conclusion:** EAGLE allows curation of evidence for association of genes with autism, as opposed to with NDDs broadly. Our analysis also revealed differential patterns of enrichment and expression profiles at the brain and cellular level, suggesting the biological relevance of differentiating ASD and the broader NDD-phenotype.

## Introduction

Clinical guidelines recommend genetic testing of individuals, usually children, with a diagnosis of autism spectrum disorder (ASD or autism)^1,2^ and genomic testing has become standard-of-care^3^. Considering rare (<1% population allele frequency, presumably penetrant) genetic variants, diagnostic detection rates in individuals having a gold-standard autism diagnosis^4^ are in the ∼15% range^5^; these proportions can increase up to 40% when selecting for people with profound autism or those with co-morbid presentations^2^. Polygenic scores for autism exist, but their explanatory power, while statistically significant, remain minimal in effect size, limiting downstream applications in research settings^6^.

There is consensus that genetic variants can contribute to autism^7,8^, but there is still little agreement on which genes are - or are not - associated with ASD liability, when impacted by a deleterious variant^9^. While the currently available lists of “ASD genes”, the shorthand for genes considered to be associated with ASD etiology, play a decisive role in both clinical and research contexts, they show substantial differences in content (Supplementary Figure 1)^9,10^. A stronger consensus is imperative because these different ASD gene lists are not only the basis for pre-clinical research, but also for the gene panels used in clinical testing.

Discrepancies between ASD gene lists can be attributed to several factors: (1) disparities in cohorts (e.g., size and ascertainment) in which they were discovered, with smaller sample sizes failing to capture the breadth of genetic heterogeneity underlying ASD; (2) differences in genome analysis technologies, with varying levels of resolution for detecting genetic variants (e.g., microarrays, exome, and genome sequencing); (3) conflating ASD with the broader category of neurodevelopmental disorders (NDDs). NDDs encompass ASD, but also other brain-related phenotypes such as intellectual disability (ID), language disorders, learning disorders, epilepsy, and attention deficit hyperactivity disorder (ADHD)^11^.

The substantial comorbidity among NDDs justifies its common use as an umbrella term, for example when recommending genetic testing and clinical consultation^12^. However, despite its high co-morbidity, ASD is also diagnosed in individuals without ID, epilepsy, or other NDDs^13^, including in people with exceptional cognitive abilities^14^. These observations indicate that the neurodevelopmental and biological differences associated with autism can exist independently of cognitive ability, and therefore, that some genes may be more related to autism, rather than to NDDs more broadly. Simply put, while every person with ASD also falls in the NDD category, many individuals with NDD do not have ASD^9^.

Large-scale genome sequencing studies, including MSSNG^5^, Simons Foundation Powering Autism Research for Knowledge (SPARK)^15^, and the Autism Sequencing Consortium (ASC)^8^, have generated lists that include a combined total of 424 unique ASD-susceptibility genes (Supplementary Figure 1). Other resources such as the Simons Foundation for Autism Research Initiative (SFARI)^16^, and the Clinical Genome Resource’s (ClinGen)^17^ have developed ASD gene lists by evaluating scientific publications to stratify genes by the level of evidence supporting their association with ASD. These efforts are comprehensive but may overestimate gene-ASD associations: SFARI’s approach lacks stringent classification guidelines, and ClinGen’s curation protocol is highly-standardized, their ID/ASD gene curation expert panels assess the relationship of genes with intellectual disability *and/or* autism, without differentiating evidence for ASD specifically^17^.

We previously developed the Evaluation of Autism Gene Link Evidence (EAGLE) curation guidelines^9^, which follow ClinGen’s gene-disease validity classification framework^17^, but considers only genetic variants in individuals with ASD (with or without ID). In doing so, EAGLE evaluates both genomic and phenotypic evidence in the peer-reviewed literature to quantitatively determine the strength of association between a gene and ASD. Additionally, high-scoring variants must be linked to cases with a confirmed ASD diagnosis, based on gold-standard diagnostic criteria (Autism Diagnostic Observation Schedule (ADOS), Autism Diagnostic Interview Revised (ADI-R), and Diagnostic and Statistical Manual of Mental Disorders (DSM))^4^.

Here, we have curated 222 genes and report their level of association with ASD, including by examining copy number and other structural variants (CNVs or SVs) of DNA, as well as mitochondrial variants using the gold-standard ASD cohorts utilized in the EAGLE process. We also assessed the frequency of occurrence of different classes of variants and their predicted genetic mechanisms of action as well or the presence of these variants in different ancestral groups, providing further evidence for pathogenicity. Among these are genes annotated as definitive by EAGLE and others with limited evidence for ASD association. The combination of an established role in NDDs and the lack of evidence for association with ASD specifically indicates that evidence for the latter set of genes is driven mainly by ID. These two gene lists, “EAGLE-definitive” and “ID-predominant” respectively, were thus obtained by juxtaposition of different curation frameworks. We subsequently, examined differences among both gene lists regarding molecular function and gene expression. We also performed expert curation of the biomedical literature to add relevant model system-and functional biology-resources that may enable preclinical research for EAGLE-definitive ASD genes.

## Materials and Methods

### EAGLE operating procedures

Detailed descriptions of EAGLE guidelines have been published elsewhere^9^. Genes of interest were identified as they became available in the scientific literature or from the SFARI-Gene project^16^. The curation process consists of a protocolized review of all available peer-reviewed publications, which are scrutinized to collect ASD-subject level data as well as experimental evidence.

Following the protocol outlined by ClinGen^18^, genomic evidence is curated by assigning scores depending on different criteria including putative mutational mechanism (see below), genotyping methods, functional variant class, inheritance pattern, and population allele frequencies. Similarly, functional evidence was also included in the assessment process and evaluated based on EAGLE’s experimental evidence summary matrix.

The final score for a gene was determined by adding the individual scores from each independent case from the case-level data component and the scores assigned from the experimental evidence component. Depending on the final score, a gene was classified into one of a six-tier classification paradigm of which the ‘definitive’, ‘moderate’ and ‘limited’ ASD genes are show in Figure 1 (the remaining three scoring categories in Table S1 are ‘strong’, ‘disputed’ and ‘refuted’).

**Figure 1.**
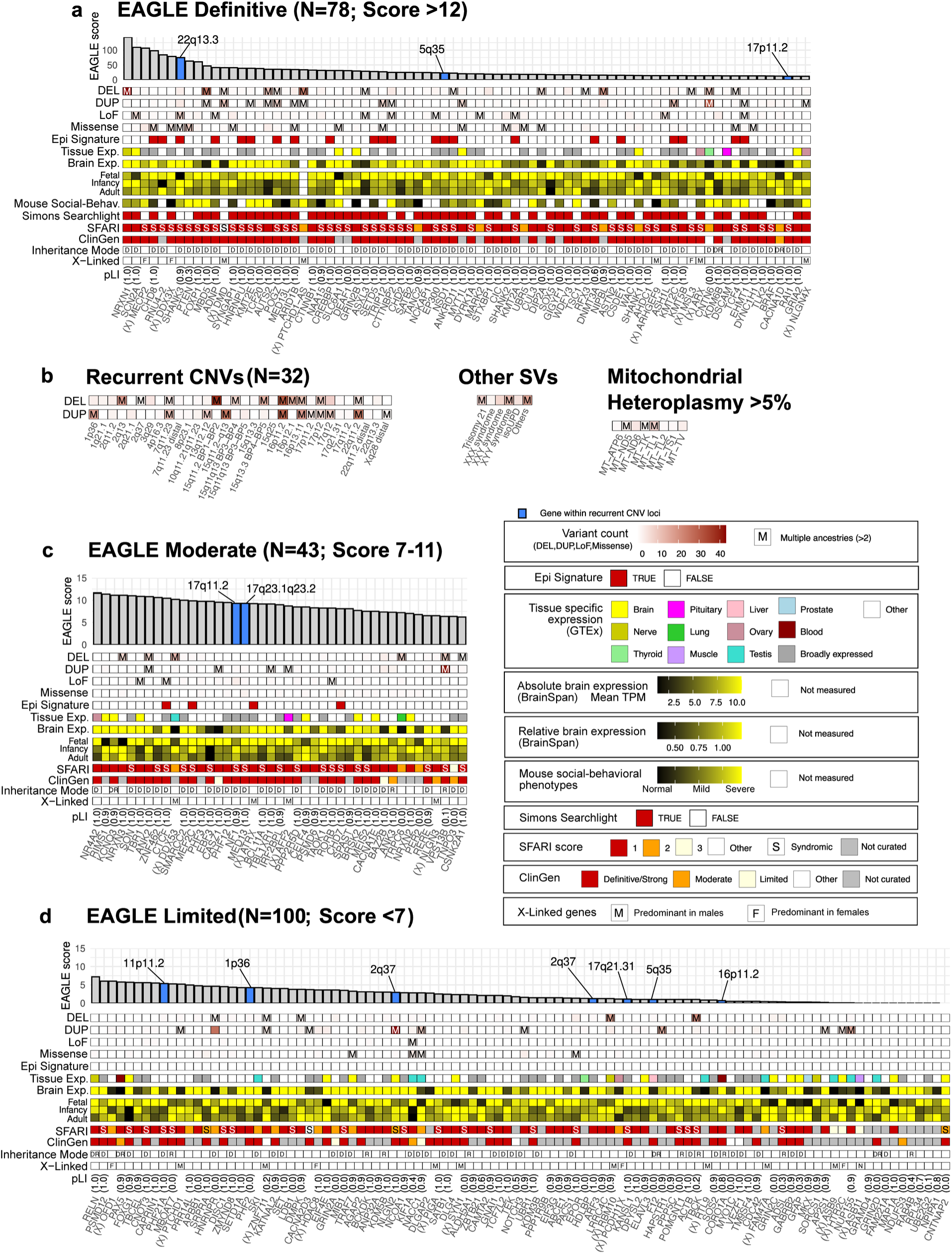
EAGLE-based compendium of genes and variants underlying ASD etiology. Manually curated genes that reached **a)** definitive **c)** moderate or **d)** limited classification, were stratified based on the EAGLE score (y-axis of the top tracks). Genes located within loci impacted by recurrent CNVs are highlighted in blue. The frequency of rare variants including CNV Deletions (DEL), duplications (DUP), loss-of-function (LoF) and missense variants was quantified among ASD cases from gold-standard phenotyped ASD cohorts (see methods). The count for each variant class is represented by an intensity gradient. Genes harboring multiple variants of a given variant class in unrelated individuals across multiple ancestries are annotated with “M”. Genes with evidence supporting a potential role of epigenetic mechanisms underlying ASD susceptibility are highlighted within the Epi Signature track. The “Brain Exp.” and “Tissue Exp.” tracks represent the expression profile of each gene within the brain and across multiple tissues, respectively. For the tissue expression track, GTEx^33^ was evaluated to determine the tissue-specific expression using a graded tissue specificity index (Tau) method^34^. If the Tau score is >0.85, it defines tissue-specific genes, while a score <0.5 defines broadly expressed genes. In the “Brain Exp.” track, the absolute brain expression level (mean TPM) of each gene was assessed using BrainSpan^26^. Additionally, the relative brain expression level across three developmental periods (Fetal, Infancy, and Adult) was calculated relative to the mean expression level across all timepoints. For the EAGLE-definitive genes only, we highlighted those reported in Simons Searchlight^35^. For autosomal genes, the inheritance pattern was annotated according to whether dominant (D) or recessive (R) mechanisms mediate the association with ASD. X-linked genes are annotated with an (X) adjacent to their gene symbol. For X-linked genes, the predominant enrichment of variants among males (M) or females (F) was highlighted. **b)** Identification of the most recurrent chromosomal abnormalities involving CNVs or larger structural variations (SVs) as well as mitochondrial heteroplasmy (>5%) implicated in ASD susceptibility. For the mouse model annotation, only germline genetic mouse models of neurodevelopmental disorders were included in the analysis. Studies employing viral induction of gene mutations in mice, as well as genetic or viral manipulation of candidate genes in other model organisms (e.g., *Drosophila melanogaster* and *Caenorhabditis elegans*), were excluded. Specific mutation target regions are annotated with exon or intron loci, together with information on cell type-specific Cre recombinase lines, point mutation sites, or targeted brain regions, when applicable. Additional relevant information can be layered on the figure as it emerges.

### Rare variant frequency in ASD

To assess the frequency of occurrence of different classes of rare variants and their putative underlying genetic mechanisms of action in EAGLE-curated genes among individuals with ASD, we analyzed one affected individual per family from MSSNG Project (4,996 individuals)^5^, SSC (3,386)^19^, and SPARK (2,419)^15^. Considering *de novo* variants, we selected only the probands with genetic information where both parents were available drawing from a total of 9,123 individuals (MSSNG:3,368, SSC:2,393 and SPARK:3,362).

We examined the 221 genes studied with the EAGLE framework (except *CNTNAP2* whose association with ASD was refuted according to EAGLE’s curation protocol) by assessing the frequency of *de novo,* rare variants in unrelated individuals with autism across genes and ancestries. These included loss-of-function (LoF) and missense single nucleotide variants (SNVs; gnomAD allele frequency <1%), small insertions and deletions (indels <50 base pairs; gnomAD allele frequency <1%), and exonic CNVs^20^ (gnomAD allele frequency <1%) affecting each gene.

### Rare variants in different ancestries

Although genetic discoveries have been largely driven by the analysis of individuals of European ancestry, overlapping genes are likely impacting ASD susceptibility across populations, which can also serve to validate pathogenicity^21,22^. To further characterize these genes, we assessed the burden of rare variants among the EAGLE-curated genes across different ancestries. To infer genetic ancestry, we assigned an individual’s genotype to the most similar ancestral reference group using principal components (PCs) computed with GCTA on the reference panel, followed by projection of target samples onto the same PC space. This procedure was performed separately for each dataset (MSSNG, SSC, and SPARK) using a population reference panel based on 266,423 consensus SNPs, following standard protocols^23^. The reference panel comprised 151 individuals from each major population group, including Africans, South Asians, East Asians, and Europeans from the 1000 Genomes Project, Middle Eastern individuals from the Human Genome Diversity Project (HGDP) and Native American individuals from the Peruvian Genome Project (PGP)^23,24^.

We then built a Random Forest classifier to predict the ancestry of each sample. The model generated prediction probabilities across the six major ancestry groups. Each sample was assigned to the ancestry group with a predicted probability >50%. Variants observed in multiple ancestry groups were labeled as “M” in the matrices shown in Figure 1.

### Definition of gene lists used for computational analyses

For the subsequent analyses performed in this study, we compared two gene lists; (1) “EAGLE-definitive” as defined in this study (n=78) and (2) “ ID-predominant” list comprising genes that reached limited classification according to EAGLE but classified as definitive or high confidence by ClinGen or SFARI-Gene (n=69). The rationale for this classification and naming scheme aligns with emerging literature recognizing data indicating differences in the relative frequency of variants among genes identified in ASD compared to ID-ascertained cohorts^8^. We postulate that the reason a gene is defined as definitive by SFARI and/or ClinGen but fails to reach definitive classification under an ASD-framework such as EAGLE is because its association is mainly driven by its relationship to ID rather than to ASD.

### Gene-set enrichment analysis

To identify biological pathways that are enriched or shared between the two gene lists, we applied gene-set enrichment analysis (Figure 2). First, GO (Gene Ontology) terms significantly enriched for each gene list were obtained from g.Profiler web tool using default parameters. For any gene with an ambiguous Ensembl identifier, the transcript with the most GO annotations was selected. A high-quality network based on functional biological information was created using the Enrichment Map application from Cytoscape (Version 3.10.3)^25^. For this analysis, we only retained gene-sets with a ‘term-size’ ranging from 100-4000 genes. The network was built using a false discovery rate (FDR) q-value cut-off of 0.01 and the edge cut-off filter (Jaccard’s index similarity) of 0.5 to reduce overlap within clusters and make the network more scattered. We used the default parameters from Enrichment Map for the remaining options.

**Figure 2.**
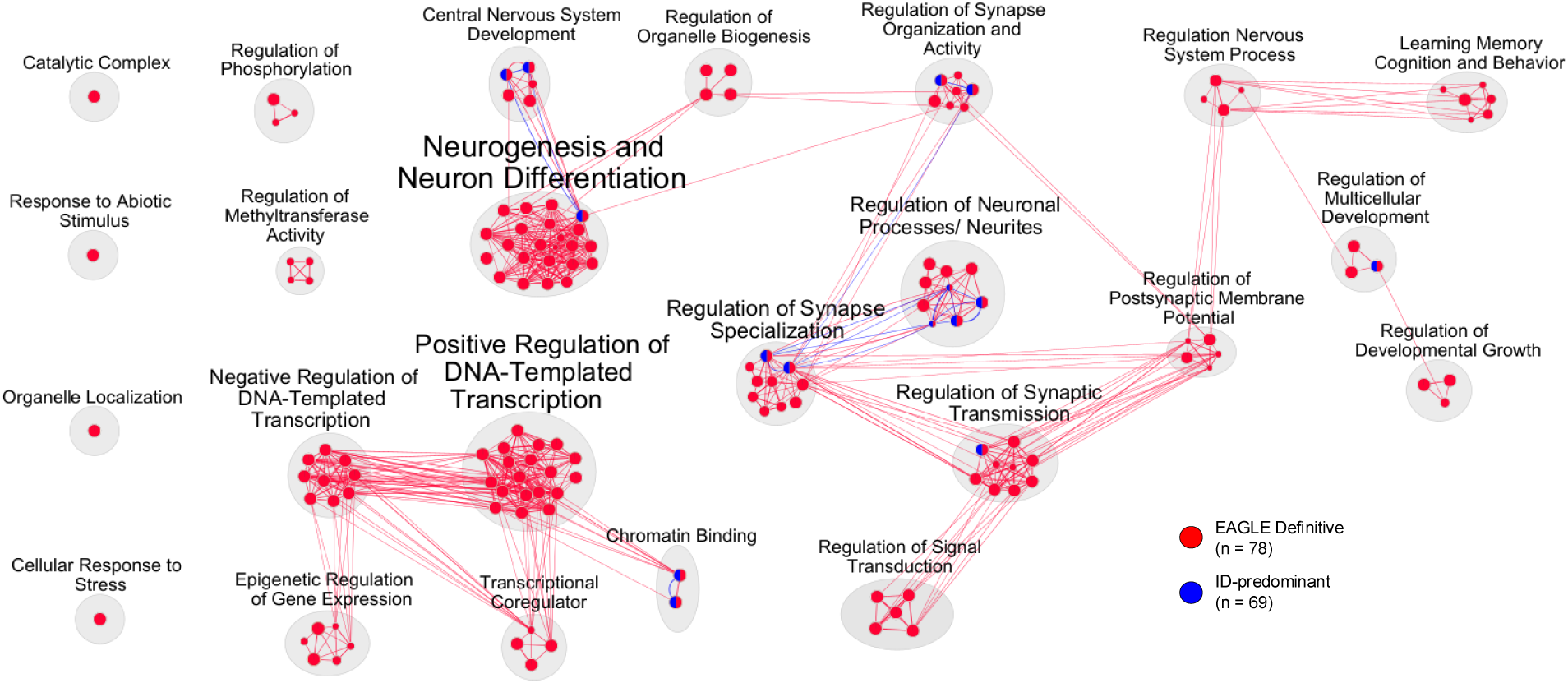
Differential enrichment of gene sets among EAGLE-definitive compared to ID-predominant genes. Each node (circle) represents a gene set that was significantly enriched among EAGLE-definitive genes (red) or ID-predominant genes (blue). Closely related gene sets were grouped into broader functional clusters (light grey).

### Brain spatiotemporal expression

Spatiotemporal gene expression data was obtained from BrainSpan, a resource that provides RNA sequencing data for 26 different cortical and subcortical structures derived from 42 brain specimens covering prenatal to adult development (Figure 3)^26^. Therefore, we grouped samples into five developmental periods (Fetal, Infancy, Childhood, Adolescence, Adulthood), including all prenatal samples grouped as fetal period. The 26 brain structures were then grouped into three distinct brain regions, which include the cortex, subcortex, and cerebellum. These brain regions and corresponding brain structures are outlined in Table S2. Then, we only retained brain structures collected in at least five independent samples across all developmental periods (resulting in 18 unique brain structures remaining). Additionally, we excluded any gene lacking corresponding EntrezGene ID. Additional details are presented in the Supplemental Methods.

**Figure 3.**
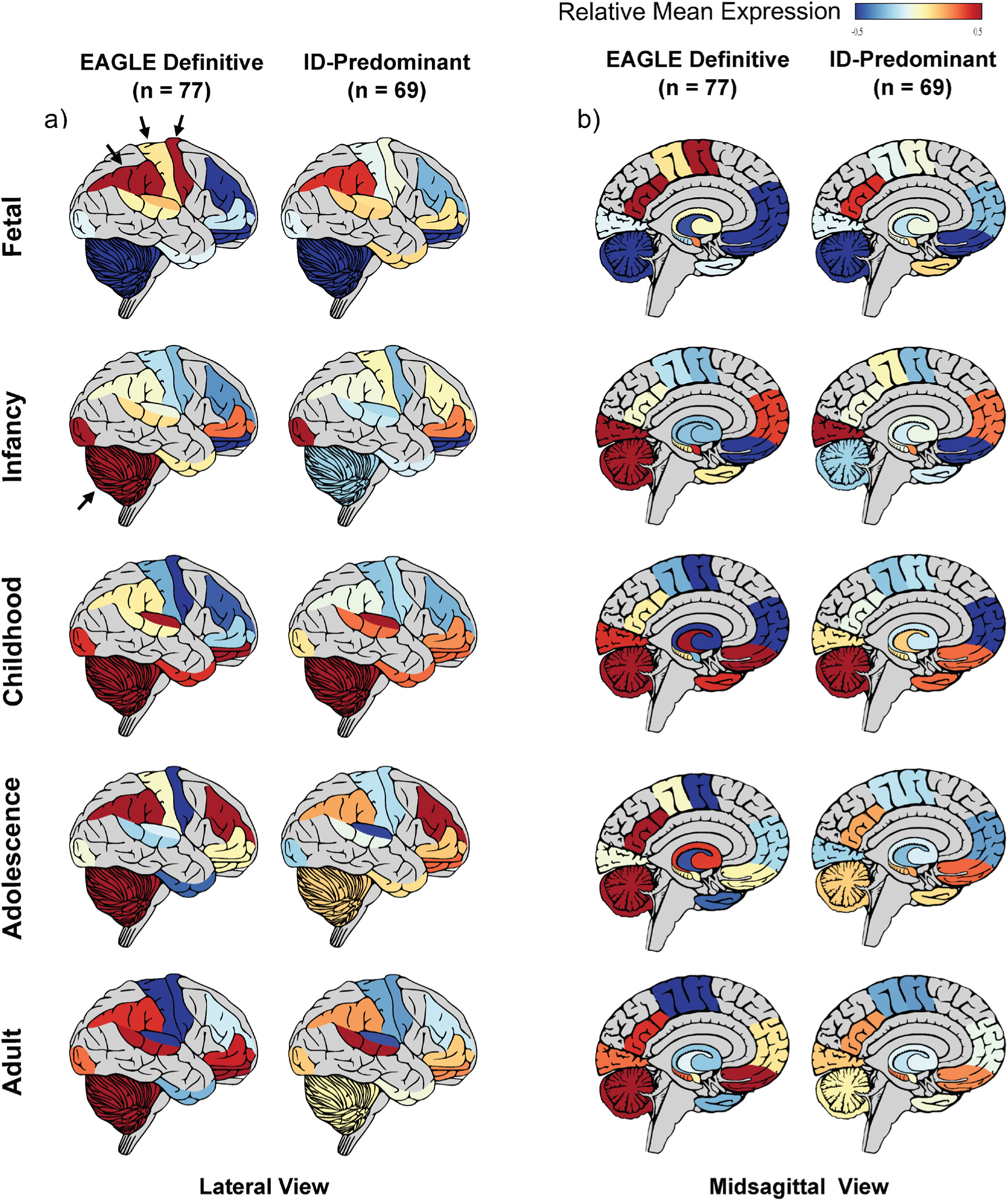
**General expression patterns across brain structures**. **a)** Lateral and **b)** midsagittal views of the human brain across five developmental periods, comparing the expression patterns of EAGLE-definitive and ID-predominant genes. Brain structures displaying over-expression (red) or under-expression (blue) of gene lists are highlighted. If expression data was not available, the brain structure was labelled in grey. For the EAGLE-definitive gene list, the lncRNA *PTCHD1-AS* was not included in the analysis since its expression was not reported in BrainSpan.

### Cell-type-specific Expression in Neuronal Subtypes

We retrieved single cell transcriptomic data from the multi-omics atlas of the developing human neocortex from The BRAIN Initiative Cell Atlas Network (BICAN)^27^. This atlas contains single-nucleus, paired RNA sequencing (RNA-seq) data from 232,328 nuclei extracted from the prefrontal cortex (PFC) and the primary visual cortex (V1), defining 33 distinct cell-types based on transcriptomic profiling (Figure 4). The nuclei were obtained from 38 human neocortical samples spanning five developmental timepoints including first, second, and third trimester, infancy, and adolescence. We normalized raw RNA-seq counts and scaled with Seurat^28^, using defined cell-type clusters for normalization. We then examined differential expression across the cell-type clusters. For this analysis, we excluded 1) genes expressed in <5 nuclei; 2) potential nuclei doublets as determined by Scrublet (v0.2.2)^29^; 3) nuclei with gene counts <3 median absolute deviations (MADs) and <300 genes; 4) nuclei with ribosomal or mitochondrial gene count percentages >20% or 3% per sample MADs, respectively; 5) cells with highly expressed *MALAT1* gene and 6) combination of cell-type and developmental timepoint with <50 cells. After quality control, we used the FindAllMarkers function within the Seurat package to calculate the log2 FC (average log-fold change) for each gene across all defined cell-type clusters. The log2 FC values were normalized using z-transformation.

**Figure 4.**
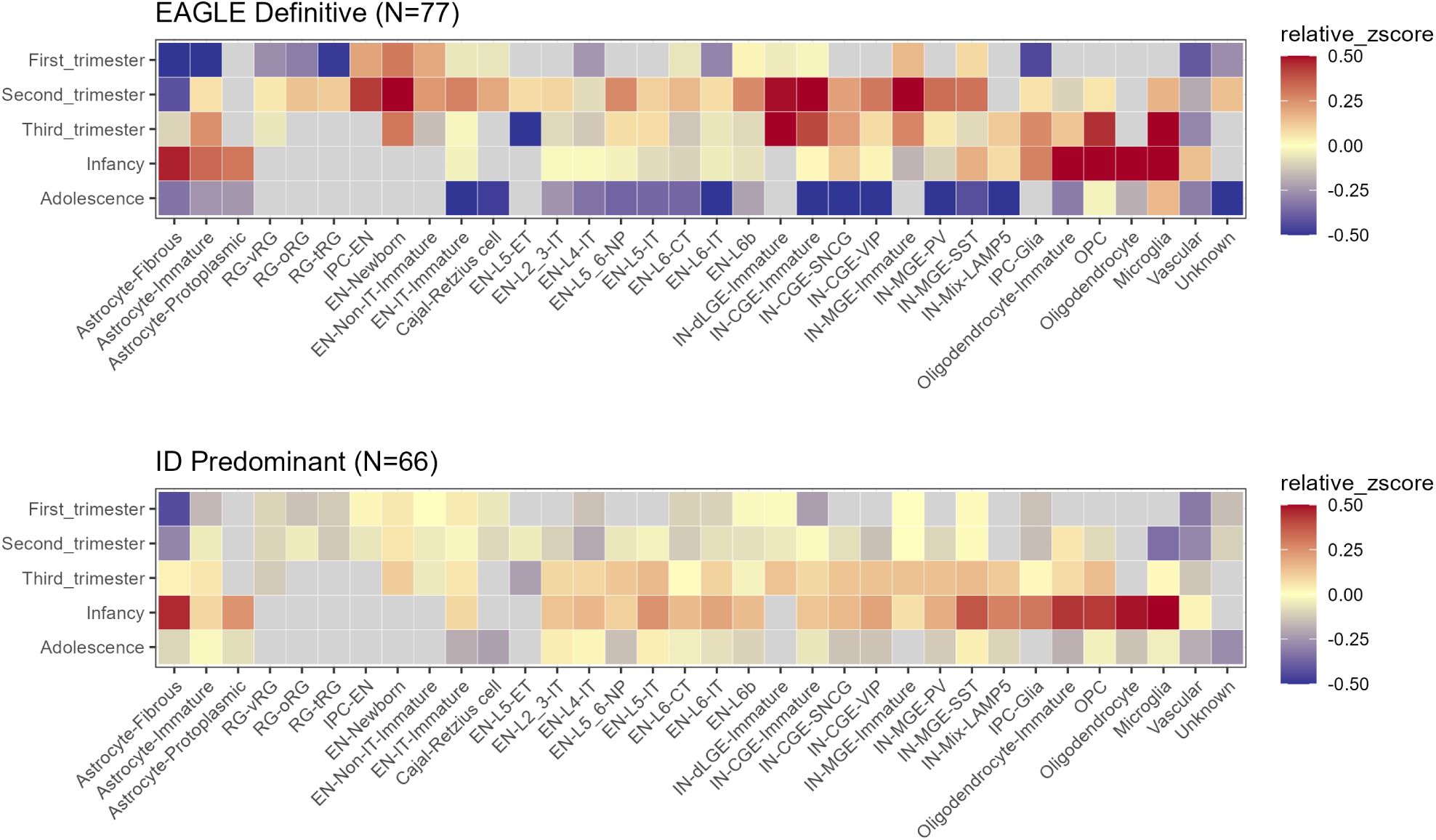
Differential expression analysis across neuronal cell-types: The expression profiles of **a)** EAGLE-definitive genes and **b)** ID-predominant genes were compared across neuronal cell-types and developmental periods. The average increased expression (red) or reduced expression (blue) across genes within each gene list was plotted relative to a baseline. To visualize the cell-type-specific trends of our gene lists (EAGLE-definitive vs. ID-predominant), we employed a relative enrichment profiling approach consistent with our brain spatiotemporal analysis. For each cell-type cluster, we calculated the mean normalized expression of the genes within our target lists. We then subtracted a background baseline, defined as the mean normalized expression of human protein-coding genes not associated with ID/ASD (n=19,708). To ensure the robustness of the observed trends, we excluded any genes expressed in less than 10% of all cell types. For the EAGLE-definitive gene list, snRNA *RNU4-2* was not included in the analysis since its expression was not reported in the BICAN Atlas. For the ID-predominant list, *PAX5* was not captured in the Atlas, while the genes *SETD1A* and *BCKDK* were excluded after quality control.

### Preclinical data curation

To help enable preclinical research, we used the 78 EAGLE-definitive genes as an anchor to curate ASD mouse models and cellular biology resources (Table S3) or link to other existing curated resources (Simons Searchlight for induced pluripotent stem cells and/or organoids) (Figure 1). Murine-models from the literature were examined for their corresponding behavioral phenotype data leading to the classification of diverse behavioral phenotypes into five categories—social interaction, social communication (ultrasonic vocalization; USV), repetitive behavior, learning and memory, and motor functions. The social interaction category includes the three-chamber social interaction assay (assessing sociability and social novelty preference), reciprocal social interaction tests, and social hierarchy tests. Mouse lines exhibiting statistically significant alterations in one or more, but not all, of these assays were classified as showing a “mild impairment” in social interaction.

The availability of protein-based research tools to enable cell biology and translational studies was collated using data from the Structural Genomics Consortium, the Protein Data Bank (PDB), UniProt, YCharOS, and associated literature. Information collected included protein identifiers, availability of experimentally determined structures, number of deposited PDB entries, structural coverage, resolved domains, construct boundaries, recombinant expression systems, expression hosts, availability of purified proteins, availability of knock-out validated renewable antibodies, and supporting publications (Table S4).

## Results

### EAGLE-annotation characteristics for ASD

EAGLE can be differentiated from other existing ASD gene curation frameworks. Compared to the ClinGen ID/ASD-GCEP^17^, EAGLE implements the same gene scoring protocol, to which a measure of confidence of the reported phenotype (ASD) is added^9^. In contrast to ClinGen’s curation protocol, EAGLE allows scores to increase based on all available evidence, even after the threshold for definitive classification (score=12) is reached (Table S1), providing significant depth in the genotype-phenotype data. The resulting continuous metric allows consideration of ASD-related genes along a gradient of evidence for association. In contrast to ClinGen’s ID/ASD approach, which considers ID and/or ASD as relevant phenotypes^17^, EAGLE restricts evidence to cases with ASD, with or without ID; compared with SFARI^16^, it also applies more stringent criteria for counting ASD-specific phenotypic evidence. EAGLE also enables the identification of definitive ASD susceptibility genes whose associations are primarily driven by inherited LoF variants or CNVs, as exemplified by *ASTN2* (EAGLE score=15.75; SFARI score 2 (Strong candidate)) or by *de novo* gain-of-function variants, as the case of *CACNA1D* (EAGLE score=12.7; SFARI score 2). The X-linked lncRNA *PTCHD1-AS* (EAGLE score=33.6) offers another relevant example of the utility of EAGLE-scoring in ASD^30^.

### ASD candidate gene testing using EAGLE

We prioritized 222 genes for EAGLE curation based on studies suggesting their potential association with ASD, or because they were included by SFARI-Gene^16^. The distribution of EAGLE scores of these 222 genes is summarized in Figure 1 and Tables S5-S6. Of all curated genes, 78 (34.6%) were categorized as definitive; one gene, *MSL3*, was classified as strong, since at the time of curation, the evidence supporting its association with ASD had not been replicated for more than 3 years; 43 (19.4%) reached moderate level of association; and 99 (44.6%) demonstrated limited evidence for a role in ASD. The association for *RELN* was disputed due to the presence of contradictory evidence; similarly, the association of *CNTNAP2* was refuted, since the evidence relies on common variants that were not replicated by larger case-controls studies^31^. Additionally, although rare damaging variants have been reported in this gene, evidence for enrichment in cases is lacking^31^. Established ASD susceptibility genes obtained the highest scores, including *NRXN1* (score=143.75), *SCN2A* (109.3), *CHD8* (97.65), *SHANK3* (74.85), and *PTEN* (63.15). The evidence for *NRXN1* was primarily driven by N-terminal deletions of the gene, which has multiple isoforms (Supplementary Figure 2a)^32^. The investigation of variant classes driving the association, provides insights into possible underlying disease mechanisms involved in ASD etiology. For example, the association with ASD of *BRAF* and *CACNA1D* appears mediated by the action of gain-of-function variants.

### Definitive ASD genes ascertained by different resources

Most genes (n=210) evaluated by EAGLE have also been curated by SFARI-Gene and/or ClinGen’s ID/ASD-GCEP. The remaining (n=12) were not yet included in these resources at the time of EAGLE curation. We examined differences in curations in overlapping genes between the three frameworks. As of October 2025, SFARI reports 240 high-confidence genes, with 171 of these overlapping with the EAGLE manually curated genes. Of these, 68 (39.8%) genes reached definitive classification according to EAGLE, while 39 (22.8%); 63 (36.8%); and 1 (0.6%) were classified as moderate, limited, or disputed respectively (Supplementary Figure 3a). Similar results are observed by analyzing SFARI’s strong candidate genes (Score 2 or 2S; analogous to EAGLE’s moderate level of association). A total of 26 genes in this category were also curated by EAGLE. Of these, 3 (11.5%) were classified as moderate by both protocols, while 13 (50%) genes were defined as limited based on EAGLE. Notably, EAGLE classified 9 (34.6%) genes as definitive (Supplementary Figure 3b).

A similar pattern was observed regarding genes curated by ClinGen’s ID/ASD-GCEP^17^. From the genes classified as definitive by the ID/ASD-GCEP, 94 were also curated by EAGLE. Of these, 49 (50.5%) genes were classified as definitive by both frameworks, while 23 (23.7%), and 25 (25.8%) were curated by EAGLE as moderate or limited, respectively (Supplementary Figure 3c).

### Rare Variant Frequency in EAGLE genes

Among the 78 EAGLE-definitive genes, 37 (47.4%) harbored variants observed across multiple ancestry groups. Some of these cross-ancestry signals were restricted to specific variant classes within each gene; examples include CNV deletions in *NRXN1*, LoF variants in *SCN2A*, missense variants in *CHD8*, and duplications in *KMT2C*, suggesting that distinct molecular mechanisms may contribute to ASD susceptibility across those genes (Figure 1-a). In contrast, some genes exhibited multiple variant classes with elevated frequency found across different ancestry groups^21^. For example, both deletions and duplications were observed for *MBD5* and *DMD*; duplications and LoF variants for *CHD2* and *EHMT1*; and both LoF and missense variants for *SHANK3*. These multi-ancestry signals were less frequent among genes with lower EAGLE evidence classifications, with 12 out of 43 (27.9%) moderate EAGLE genes and 17 out of 99 (17%) limited EAGLE genes showing variants across multiple ancestry groups (Figure 1a).

### X-linked genes and estimates of sex difference in disorder prevalence

Difference in prevalence between males and females having ASD is typically ∼4:1^36^, implicating a role for the sex chromosomes^36^. As part of the EAGLE curation protocol, we recorded patients’ sex and calculated separate scores for males and females. Collectively, for the 25 X-linked genes that were evaluated by EAGLE, 381 males and 299 females were included in the case-level component of evidence assessment. Eight genes reached a definitive-level of association, while four and thirteen genes were classified as moderate and limited by EAGLE, respectively. The availability of these data enabled us to identify genes that are predominantly altered in one biological sex, such as *MECP2*, *DDX3X* and *MSL3* predominantly impacting females (Figure 1). To determine whether the number of males and females that were included in the EAGLE’s evidence assessment process also reflect the male:female (M:F) diagnosis ratio for ASD, we compared this ratio among different gene lists. First, we calculated the M:F ratio among the 10 EAGLE-definitive and moderate X-linked genes. We excluded *MECP2* (males=49; females=133) and *DDX3X* (males=14; female=109) since both genes are responsible for X-linked dominant disorders primarily found in females, due to high lethality in male carriers. In this gene list, the M:F ratio was 6.89 (262 males:38 females; p-value= 7.339e-4; two-sided Wilcoxon rank-sum test) representing a significant difference in diagnostic rate among males and females (Supplementary Figure 4). Then, we calculated the diagnostic ratio among the EAGLE-definitive genes (n=70; excluding the eight definitive X-linked genes). The M:F ratio was 2.96 (2,247 males:758 females; p-value= 2.367e-11; two-sided Wilcoxon rank-sum test). Among the ID-predominant autosomal genes (n=64), the ratio was 4.05 (300 males:74 females; p-value = 3.253e-12; two-sided Wilcoxon rank-sum test).

### Enrichment of distinct genes sets relevant to ASD

Gene set enrichment analysis allowed us to compare biological functions that are similarly or differentially enriched among the genes of the two gene lists of interest (Table S6). We observed 24 gene-set clusters enriched for the two gene lists and 16 of them were exclusively enriched for EAGLE-definitive genes. These clusters ranged from groupings of “regulation of DNA-templated transcription”, “epigenetic regulation of gene expression”, “regulation of methyltransferase activity”, “learning, memory, cognition and behavior”, and “regulation of phosphorylation” (Figure 2). Gene-set clusters associated with “neurogenesis and neuron differentiation”, “central nervous system development”, “regulation of synapse specialization and neuronal processes”, as well as “chromatin binding” were enriched in both, EAGLE-definitive and ID-predominant gene lists. We did not observe any functional cluster that was exclusively enriched in the ID-predominant genes. To examine whether these differential enrichments were not driven solely by difference in gene lists size, we compared the enrichment level of all 141 enriched gene sets using odds ratio (OR) at gene set-level, as well as, at gene-set cluster level (mean OR). We observed no gene sets with lower enrichment level in EAGLE-definitive compared to ID-predominant genes (Supplementary Figure 5a), suggesting that EAGLE-definitive genes converge on highly overlapping molecular functions and cellular processes, while ID-predominant genes are functioning on broader biological pathways. Notably, the most distinct differences were observed for clusters associated with “learning, memory, cognition, and behavior”, “epigenetic regulation of gene expression”, “regulation of methyltransferase activity” and “regulation of phosphorylation”.

To further characterize the enriched clusters and determine whether their aggregate enrichment was driven solely by a subset of the EAGLE-definitive genes, we mapped the membership of each of the EAGLE-definitive genes among the 24 gene-sets clusters (Supplementary Figure 5c). We identified three major subgroups enriched for most EAGLE-definitive genes. These include a subgroup related to regulation of gene expression (Negative and Positive regulation of DNA-Templated transcription, and chromatin binding), a second subgroup implicated in early developmental process of central nervous system development (e.g., neurogenesis) and a third one associated with late neurodevelopmental processes including synapse specialization, synaptic transmission and neurites growth.

### Characterization of brain spatiotemporal differential expression

To identify biological insights at the spatiotemporal expression level in the brain among EAGLE-definitive and ID-predominant genes, we visualized their expression profile across 18 brain structures (Table S2) along the trajectory of human brain development^26^. This approach allowed us to highlight general expression patterns within brain structures for the two gene lists (standardized mean expression values for each gene list were plotted relative to a baseline). We observed higher expression levels among the EAGLE-definitive genes relative to the ID-predominant genes, particularly during prenatal development (Figure 3). During this developmental period the primary motor cortex (M1C), primary somatosensory cortex (S1C), and inferior parietal cortex (IPC) showed consistently higher expression among EAGLE-definitive genes (Figure 3 and Supplementary Figure 6).

In the post-natal period, the most interesting finding is represented by the consistently high relative expression of EAGLE-definitive genes within the cerebellum across all four post-natal stages. In particular, the expression signature of EAGLE-definitive genes is considerably higher than ID-predominant genes in the cerebellum during infancy.

### Cell-type-specific Expression Profiling in Neuronal Subtypes

To map the distribution of EAGLE-curated genes across neuronal cell-types and developmental timepoints, we utilized the high-resolution BICAN single-cell atlas^27^. By comparing the relative mean expression patterns of EAGLE-definitive genes and ID-predominant genes, we identified cell types that showed relatively higher expression level during specific developmental periods. Specifically, during the second trimester, intermediate progenitor cells of excitatory neurons (IPC-EN) and glutamatergic EN-newborn cells showed differences between the two gene lists. To a lesser extent, immature intratelencephalic (IT) and non-intratelencephalic (non-IT) excitatory neurons also exhibit increased expression only among EAGLE-definitive genes. Certain subtypes of inhibitory neurons (INs), including immature INs within the dorsal lateral ganglionic eminence (IN-dLGE) and the caudal ganglionic eminence (IN-CGE) showed higher expression levels among EAGLE-definitive compared to ID-predominant genes. Finally, during the third trimester, both oligodendrocyte precursor cells (OPCs) and microglia display the most variable expression differences amongst gene lists.

### Behavioral Phenotypes in Murine Preclinical Models

To characterize autism-related behavioral phenotypes in mouse models, we compiled behavioral data from the published literature and generated a summary table of mouse models carrying mutations in EAGLE definitive genes (Figure 1a; Table S3).

Assessment of social interaction is a primary approach for evaluating autism-related behavioral phenotypes in mice. Except for *Ddx3x* and *Rnu4-2*, 8 of the top 10 EAGLE-ranked genes recapitulated abnormalities in social interaction behavior. Many of these models exhibited partial impairments, showing deficits in one or more, but not all, social interaction assays. For example, mouse models carrying mutations in *Nrxn1α*, *Scn2a*, *Chd8*, or *Shank3* displayed normal sociability in relatively simple social interaction paradigms but showed impairments in more demanding tasks assessing social novelty preference. Furthermore, mutations affecting distinct isoforms within the same gene (e.g., *Nrxn1α* versus *Nrxn1β*, *Chd8S/L* versus *Chd8L*, and *Shank3a* versus *Shank3b*) often produced divergent autism-related behavioral phenotypes.

The use of cell type-specific Cre recombinase lines further revealed differential contributions of distinct neuronal populations to autism-related behaviors. For example, *Mecp2* deletion in glutamatergic neurons or parvalbumin-positive interneurons resulted in social behavioral abnormalities, whereas deletion in somatostatin-positive interneurons largely preserved typical social behavior.

We also found that relatively limited data are available regarding ultrasonic vocalizations (USVs) in mouse models of autism, despite growing interest in this area driven by recent advances in machine learning-based analyses of mouse communication^30^. Finally, although most mouse models carry mutations targeting specific exons, several studies have generated models harboring CNV deletions, duplications, LoF variants, or missense mutations. These models provide valuable opportunities to examine genotype–phenotype relationships with greater precision.

## Discussion

We curated 222 genes previously reported as associated with NDDs broadly, resulting in 78 genes with EAGLE-definitive, 43 with moderate and 99 with limited evidence for association with ASD. Comparing EAGLE-definitive to ID-predominant gene-sets, we observe distinct biological signatures, as demonstrated by significant differences in gene-set enrichment, brain and cell type-specific expression patterns. Additionally, we summarized 32 recurrent CNVs and 14 structural and mitochondrial variants regularly found to be pathogenic in autism (Figure 1), and also present variant classes (e.g., LoF, missense, duplication, deletion, epi-modification) and sites of alteration, along with occurrence of variants across different ancestries, in 37 of the 78 genes for which such data were available (noting there is still paucity of whole genome sequence data in ASD from non-European populations). *SHANK3*, a prototypical ASD gene (EAGLE score 74.85)^37^ harbors all classes of variants (Supplementary Figure 2) impacting ASD susceptibility, with some occurring recurrently at the same nucleotide site in unrelated families from different ancestries^22^.

A possible limitation of EAGLE is that more recently described autism-related genes, which have fewer observations, would yield lower scores. Consequently, we anticipate that some of the 43 EAGLE-moderate genes will likely move into the EAGLE-definitive category when future studies present additional evidence (e.g., *DDX53*)^38^. A total of 99 genes were classified as EAGLE-limited; while all genes in this category are involved in the broader category of NDDs, current evidence for a link with ASD specifically was limited for this subset. While new evidence in support of an etiologic role in ASD may transpire in the future, we submit that the current lack of evidence suggests that these genes should not be considered as “ASD genes”.

The availability of a ‘gold-standard’ gene list for autism should also help to re-affirm true etiology from increasingly pervasive misinformation^39^. To make our results accessible to clinical geneticists and fundamental researchers considering pre-clinical models, we present our data in Figure 1 in a ‘periodic-table style’ with structured elements amenable for continual annotation. EAGLE-gene annotations are regularly updated and posted online (https://gene.sfari.org and http://eagle.tcag.ca/app). We have also developed a large-language model workflow for automated extraction and scoring of literature linking genes to ASD, which have increased ongoing annotation efficiencies^40^.

Numerous studies have shown that ASD often co-occurs with other NDDs and psychiatric conditions^41^, implicating common molecular and cellular origins. This may be particularly pertinent to ASD and ID, as an estimated 45% of individuals with ASD also have ID^41^. The EAGLE framework acknowledges that despite the frequent comorbidity of ASD and ID, both phenotypes also occur independently. Whether some genes may be predominantly involved in ASD, as opposed to a scenario where any involved gene is always equally related to both ASD and ID, is subject to ongoing academic discussion^42,43^.

The differential patterns we observed across gene-set enrichment, whole-brain expression, and cell-type-specific expression data provide strong biological support for a set of genes predominantly involved in ASD (Figures 2–4). At the molecular level we identified differential enrichment of gene-sets relevant to ASD genes, while no functional cluster was exclusively enriched for ID-predominant genes.

Characterizing brain expression levels, we detected regions with high relative expression of EAGLE-definitive genes including the primary motor cortex during fetal development and the cerebellum in infancy. This is consistent with a growing body of evidence suggesting that dysfunction of cerebellar systems is underlying altered motor phenotypes including excessive stereotyped and repetitive behaviors^44^. Similarly, at the cellular level, we observed relative increased expression of ASD genes in excitatory neuronal cell types. These results are highly consistent with previous findings showing that disruption of cortical circuits targeting glutamatergic neurons more likely affect core ASD phenotypes including social domains^45^.

Overall, these findings are also consistent with a growing body of genotype–phenotype studies showing that some genes are more specifically associated with ASD, whereas others are more broadly linked to NDDs^30,46^. The findings are important not only for their clinical implications, but also for their mechanistic implications. The fact that ASD gene lists, including EAGLE, guide genetic testing^3,47^, support preclinical model development, inform emerging target-based therapeutic strategies including the search for renewable antibodies targeting ASD-associated proteins (Table S4), and guide clinical trials^48,49^, underscore the ongoing need for precision in the curation of gene associations with autism.

### Data Availability Statement

The data generated and analyzed during this study are included in the published article and its supplementary material. EAGLE curation files for individual genes are available upon request from the corresponding authors. EAGLE-gene curations are regularly updated and can be found online (https://gene.sfari.org and http://eagle.tcag.ca/app).

## Acknowledgements

We wish to acknowledge the following resources: MSSNG (www.mss.ng), by Autism Speaks and The Centre for Applied Genomics at The Hospital for Sick Children, Toronto, Canada; and SPARK (www.sparkforautism.org), by the Simons Foundation Autism Research Initiative. We thank the participating families for their time and contributions to these databases, as well as the generosity of the donors who supported these programs. We acknowledge the Peruvian Genome Project (PGP) team for generating the dataset used exclusively as a reference panel for ancestry inference in this study. Additionally, we acknowledge Dr. Shreejoy Tripathy and Jarryll Uy for the insightful feedback provided during the editing and refinement stages of the manuscript.

## Author Contributions

J.A.S.V., and S.W.S. conceptualized and developed the EAGLE framework as part of an international, multidisciplinary team. N.B.S., and O.R. carried out the manual curation of the 222 genes through the implementation of the EAGLE guidelines. W.E. processed and analyzed gene expression data from BrainSpan and The BRAIN Initiative Cell Atlas Network (BICAN). M.M.A. performed the rare variant analysis in EAGLE curated genes. K.T-B., K.B. and K.B.M. performed protein annotation; C.L., and A.E., performed the search of antibodies targeting the EAGLE-definitive genes. N.B.S., J.M., J.A.S.V. and S.W.S. wrote the manuscript. N.B.S., O.R., W.E., M.M.A., J.M., V.F., X.Z., N.R.A., N.H., S.Y.K., K.S., J.A.S.V., and S.W.S significantly contributed to discussions to the content of the manuscript. N.B.S., W.E., M.M.A., J.M., J.L.H., K.S., J.A.S.V., and S.W.S reviewed and/or edited the manuscript before submission.

## Ethics Declaration

Ethical review and approval were obtained from The Hospital for Sick Children Research Ethics Board (REB# 1000080561).

## Code availability

The code used to process raw data and analyze the information is available in the repository at https://github.com/XavierBauti3/EAGLE_Manuscript.

