## Supplemental Material for "Genes/variants for diagnostic testing and pre-clinical research in autism spectrum disorder"

### Supplementary Methods

### Supplementary Tables

**Table S1.** EAGLE curated genes grouped by levels of association

**Table S2.** Categorization of brain structures into three distinct brain regions

**Table S3**. Summary of behavioral phenotypes in mouse models of ASD carrying mutations in EAGLE definitive genes.

**Table S4**. Integrated prioritization framework for autism spectrum disorder (ASD)-associated genes based on genetic evidence, structural tractability, experimental resources, and chemical biology readiness.
[Provided as a separate excel table]

**Table S5.** EAGLE annotation of the 222 manually curated genes
[Provided as a separate excel table]

**Table S6.** Genes included in the EAGLE-definitive and ID-predominant gene lists
[Provided as a separate excel table]

### Supplementary Figures

**Figure S1.** Overlap of ASD gene lists from large scale genome sequencing resources.

**Figure S2**. Detailed characterization of the allelic variation underlying the evidence supporting the association of EAGLE genes with ASD.

**Figure S3.** Discrepancies in the level of gene-ASD associations reported by SFARI-Gene, ClinGen and EAGLE.

**Figure S4.** Proportion of male and female cases included in the case-level evidence assessment process.

**Figure S5.** Differential enrichment of gene sets among EAGLE-Definitive and ID-predominant gene lists.

**Figure S6.** Reference map of brain structures.

### Supplementary Methods

#### *Brain spatiotemporal expression*

The original expression data from BrainSpan was further analyzed to identify samples and genes whose expression might represent potential outliers in the dataset. First, we carried out PCA-based outlier detection using a multivariate standard deviation approach to identify samples whose expression profile deviates by more than three standard deviations from the mean value of the corresponding first ten principal components (PC1-PC10). These samples were excluded, leaving 471 samples for further analysis. Then, to correct from potential batch artifacts, we carried out gene-wise linear regression to model donor-specific effects and obtain the residual expression values after donor effect correction. Additionally, we removed any gene for which the standard deviation of its residual expression was zero across the remaining samples, resulting in 21,051 genes retained for further analysis.

Significant inter-regional variation across the three brain regions (cerebral cortex, subcortex, and cerebellum) has been reported[1]. To avoid attenuation of expression signals between brain structures due to the inter-regional variation, we normalized the expression data for each brain region separately instead of implementing global normalization across all brain structures. To this end, the expression profile of each gene was averaged across samples for each combination of brain structure and developmental period. We reiterated this analysis for 18 brain structures and 5 developmental periods. Then, we normalized the mean residual expression values using z-transformation across the brain structures within the cortex and subcortex separately. In the absence of subregions, the cerebellum was standardized relative to the cortex and subcortex. To generate hypotheses regarding the spatial enrichment of our two gene lists (EAGLE-definitive vs ID-predominant), we plotted the normalized mean expression values among the genes in each gene list relative to a baseline reference. We determined this baseline by calculating the normalized mean expression from the remaining human protein coding genes lacking evidence supporting a role in ID and/or ASD etiology (n=19,708 genes). To this end, we identified human protein coding genes that have not been reported by SFARI-Gene and/or the ID/ASD-GCEP from ClinGen (we acknowledge potential limitation of this strategy since some genes in the baseline reference panel could still be contributing to ASD susceptibility, but current evidence is still required). Then, the relative normalized mean expression values were obtained by subtracting the baseline expression level from the normalized mean expression values of each gene list respectively. Finally, the expression values for the EAGLE-definitive and ID-predominant gene lists were plotted using the cerebroViz package[2].

***Cellular and Molecular Biology Resources***

Protein target-based cell and molecular studies, as well as translational studies, depend on high-quality research tools.  EAGLE-definitive genes were subjected to a multidimensional annotation workflow to identify high-quality genetic, structural, experimental, and chemical biology information.

Structural and protein production data were compiled from the Structural Genomics Consortium (SGC), the Protein Data Bank (PDB), UniProt, and associated literature. Information collected included protein identifiers, availability of experimentally determined structures, number of deposited PDB entries, structural coverage, resolved domains, construct boundaries, recombinant expression systems, expression hosts, availability of purified proteins, and supporting publications.

To evaluate experimental tractability, information regarding the availability of validated research reagents was collected from public repositories and commercial suppliers. This category included the availability of commercially-available recombinant antibodies (as per CiteAb.com), knockout (KO)-validated antibodies (as per CiteAb.com and ycharos.com), KO cell lines (Revvity HAP1 KO lines and Abcam catalogues), and evidence of target expression in cultured cell lines (as per DepMap.org). The presence of validated reagents was considered an important criterion supporting future cellular, biochemical, mechanistic, and translational studies.

Information regarding known ligands, chemical probes, and small-molecule modulators was collected from SGC resources and complementary public databases. The availability of chemical tools was used as an indicator of target tractability for chemical biology studies and provided an initial assessment of druggability and potential for ligand discovery campaigns.

A final prioritization step was performed by integrating information from all annotation categories. Genes were classified according to their combined genetic relevance, structural tractability, availability of experimental resources, and suitability for chemical biology applications. Protein targets displaying favorable characteristics across multiple categories were considered high-priority candidates for recombinant protein expression, purification, structural characterization, functional assays, and screening campaigns aimed at identifying novel chemical probes and potential therapeutic modulators associated with ASD-related molecular pathways.

**Methods References**


1 Li M, Santpere G, Imamura Kawasawa Y, Evgrafov OV, Gulden FO, Pochareddy S *et al.* Integrative functional genomic analysis of human brain development and neuropsychiatric risks. *Science*. 2018;362 https://doi.org/10.1126/science.aat7615

2 Bahl E, Koomar T, Michaelson JJ. cerebroViz: an R package for anatomical visualization of spatiotemporal brain data. *Bioinformatics*. 2017;33:762–763. https://doi.org/10.1093/bioinformatics/btw726

### Supplementary Tables

#### Table S1. EAGLE curated genes grouped by levels of association

| EAGLE Classification | Score | Number of Genes |
|---|---|---|
| Definitive | 12+<br>& replication over time | 77 |
| Strong | 12+ | 1 |
| Moderate | 7-11 | 43 |
| Limited/ No Scorable Evidence* | 0-6 | 99 |
| Disputed† | NA | 1 |
| Refuted† | NA | 1 |

*Note: The evidence supporting the association of a gene with ASD is determined as non-scorable when rigorous assessment of the genotyping and phenotypic evidence results in an overall score of zero. This represents genes with limited number of cases reported in the literature, or low confidence in the phenotypic quality reporting the ASD diagnosis. † For the disputed classification, contradictory evidence does not outweigh evidence supporting a potential role in ASD, while refuting evidence significantly outweighs existing supporting evidence

**Table S2. Categorization of brain structures into three distinct brain regions**

| Cortex | Subcortex | Cerebellum |
|---|---|---|
| Primary motor-sensory cortex | Amygdaloid complex | Cerebellum |
| Posterior (caudal) superior temporal cortex (area 22c) | Dorsal thalamus | |
| Anterior (rostral) cingulate (medial prefrontal) cortex | Mediodorsal nucleus of thalamus | |
| Dorsolateral prefrontal cortex | Hippocampus (hippocampal formation) | |
| Orbital frontal cortex | Striatum | |
| Inferolateral temporal cortex | | |
| Ventrolateral prefrontal cortex | | |
| Primary auditory cortex (core) | | |
| Primary visual cortex (striate cortex, area V1/17) | | |
| Primary motor cortex (area M1, area 4) | | |
| Posteroventral (inferior) parietal cortex | | |
| Primary somatosensory cortex (area S1, areas 3,1,2) | | |

**Table S3**. Summary of behavioral phenotypes in mouse models of ASD carrying mutations in EAGLE definitive genes.

| **Gene** (cytoband) | **ClinGen curation** - SFARI score - EAGLE score | **Mouse models** | **Social interaction** | **Social communication (Ultrasonic vocalization)** | **Repetitive behavior** | **Learning and memory** | **Motor functions** | **Ref** |
|---|---|---|---|---|---|---|---|---|
| ***NRXN1*** (2q16.3) | Complex NDD - SFARI: 1 - EAGLE: 143.75 | Nrxn1α (Δexon1) | Mild impairment* | Impaired (pup calls) | Impaired | Impaired* | Enhanced | Etherton et al., 2009; Grayton et al., 2013; Dachtler et al., 2015; Armstrong et al., 2020; Xu et al., 2023 |
| | | Nrxn1α (Δexon9/intron17) | Mild impairment (novelty ↓) | NA | NA | NA | Enhanced | Xu et al., 2023 |
| | | Nrxn1βΔC (CaMKII-insertion) | Impaired | NA | Impaired | Normal | Normal | Rabaneda et al., 2014 |
| ***SCN2A*** (2q24.3) | Complex NDD - SFARI: 1 - EAGLE: 109.3 | Scn2a (Δexon1) | Mild impairment | Impaired | Impaired | Impaired | Hyperactivity (Anxiolytic) | Middleton et al., 2018; Tatsukawa et al., 2019; Lena et al., 2019 |
| | | Scn2a (Δexon4-6) | Mild impairment (novelty ↓) | Normal | Normal | Impaired | Hypoactivity | Shin et al., 2019 |
| | | $Scn2a^{T1898Nfs*27}$ (Δexon27) | Impaired (interaction↑) | NA | Normal | NA | Hyperactivity (Anxiolytic) | Wang et al., 2021 |
| ***MECP2*** (Xq28) | Rett syndrome - SFARI: 1S - EAGLE: 106.65 | MeCP2 KO (Δexon3-4) | Impaired | NA | Normal | Normal | Hypoactivity (Anxiety↑) | Guy et al., 2001; Shahbazian et al. 2002 |
| | | Glutamatergic KO (CaMKII-Cre KO) | Impaired | NA | NA | Impaired | Impaired (Anxiety↑) | Gemelli et al., 2006 |
| | | GABAergic KO (Viaat-Cre KO) | Impaired (interaction↑) | NA | Impaired | Impaired | Impaired | Chao et al., 2010 |
| | | (PV-Cre KO) | Impaired (interaction↑) | NA | Normal | Impaired | Impaired | Ito-Ishida et al., 2015 |
| | | (SOM-Cre KO) | Normal | NA | Impaired | Normal | Normal | |
| | | MeCP2 duplication | Impaired | NA | NA | Impaired | Hypoactivity (Anxiety↑) | Sztainberg et al., 2015 |
| ***CHD8*** (14q11.2) | Complex NDD - SFARI: 1S - EAGLE: 97.65 | Chd8ΔSL (Δexon1) | Mild impairment (novelty ↓) | NA | Normal | Enhanced | Hypoactivity (Anxiety↑) | Katayama et al., 2016; Platt et al., 2017 |
| | | Chd8 KO (Δexon5) | Normal | Normal | Normal | Impaired | Normal | Gompers et al., 2017 |
| | | Chd8ΔL (Δexon11-13) | Mild impairment (novelty ↓) | NA | Normal | Mildly impaired | Mild impairment (anxiety↑) | Katayama et al., 2016 |
| | | $\mathit{Chd8}^{N2373Kfs*2}$ (Δexon36) | Normal | Normal (pup call: impaired) | Impaired | Normal | Hypoactivity (anxiety↑) | Jung et al., 2018 |
| ***RNU4-2*** (12q24.23) | NDD with hypotonia, brain anomalies, distinctive facies, and absent language - SFARI: 1S - EAGLE: | NA | NA | NA | NA | NA | NA | NA |

| | | | | | | | | |
|---|---|---|---|---|---|---|---|---|
| ***DDX3X*** (Xp11.4) | X-linked syndromic intellectual disability<br>- SFARI: 1S<br>- EAGLE: 78.6 | Ddx3x (Sox2-Cre KO/ Δexon 2) | Normal | NA | Impaired | Mild impairment | Impaired motor coordination<br>Hyperactivity<br>Anxiety | Boitnott et al., 2021 |
| ***SHANK3*** (22q13.33) | Phelan-McDermid syndrome<br>- SFARI: 1S<br>- EAGLE: 74.85 | Shank3 KO (Δexon4-22) | Impaired | Impaired | Impaired | Impaired | Impaired | Wang et al., 2016 |
| | | Shank3a (Δexon4-9) | Impaired | Impaired | Impaired | Impaired | Impaired | Bozdagi et al., 2010; Wang et al., 2011; Jaramillo et al., 2016 |
| | | Shank3a (Δexon4-7) | Mild impairment (novelty ↓) | NA | NA | NA | NA | Peca et al., 2011 |
| | | Shank3b (Δexon13-16) | Impaired | NA | Impaired | Normal | Normal (anxiety↑) | Peca et al., 2011; Mei et al., 2016 |
| | | Shank3ΔC/ Shank3$^{G}$ (Δexon21) | Impaired/ Normal | NA | Impaired | Mild impairment | Normal | Kouser et al., 2013; Duffney et al., 2015; Speed et al., 2015 |
| ***PTEN*** (10q23.31) | PTEN hamartoma tumor syndrome<br>- SFARI: 1<br>- EAGLE: 63.15 | Pten (Δexon 4-5; Nse-Cre KO) | Impaired | NA | NA | Impaired | Hyperactivity<br>Impaired sensorimotor gating<br>Anxiety | Kwon et al., 2006; Zhou et al., 2009 |
| | | (Δexon 5) | Impaired | NA | Impaired | Normal | Normal motor coordination<br>Depression | Clipperton-Allen et al., 2014 |
| ***FOXP1*** (3p13) | Intellectual disability-severe speech delay-mild dysmorphism syndrome<br>- SFARI: 1<br>- EAGLE: 60.45 | Foxp1 (Nestin-Cre KO) | Impaired | NA | Impaired | Impaired | Hyperactivity<br>Impaired sensorimotor gating | Bacon et al., 2015 |
| | | Foxp1 (Emx1-Cre KO) | Impaired | Impaired | NA | Impaired | Hyperactivity<br>Anxiety | Araujo et al., 2017 |
| ***MBD5*** (2q23.1) | Complex NDD<br>- SFARI: 1S<br>- EAGLE: | Mbd5 (gene-Trap insertion in intron 2) | Impaired (interaction↑) | NA | Impaired | Impaired | Impaired motor coordination and grip strength | Camarena et al., 2014 |
| ***ADNP*** (20q13.13) | ADNP-related multiple congenital anomalies-intellectual disability-ASD<br>- SFARI: 1S<br>- EAGLE: 41.5 | Adnp (Δexon 3-5) | Mild impairment | Impaired | NA | Impaired | Impaired motor coordination | Amram et al., 2016; Hacochen-Kleiman et al., 2018 |
| ***DMD*** (Xp21.2-p21.1) | Progressive muscular dystrophy<br>- SFARI: S<br>- EAGLE: 40.95 | Dmd (Dmd$^{Q995*}$) | NA | NA | NA | Enhanced | Defensive behavior ↑ | Sekiguchi et al., 2009 |
| | | Dp71 (Dp71-specific Δexon 1) | NA | NA | NA | Impaired | Hyperactivity | Daoud et al., 2009 |
| ***SYNGAP1*** (6p21.32) | Complex NDD<br>- SFARI: 1S<br>- EAGLE: 40.75 | Syngap1 (Δexon 6-7) | Mild impairment (social novelty) | NA | Impaired | Impaired | Hyperactivity<br>Anxiolytic<br>Seizure<br>Impaired sensorimotor gating | Guo et al., 2009; Clement et al., 2012 |
| | | Syngap1 (Δexon 4-9) | NA | NA | NA | Impaired | Hyperactivity<br>Anxiolytic | Muhia et al., 2010 |
| ***HNRNPU*** (1q44) | Complex NDD<br>- SFARI: 1S<br>- EAGLE: 38.8 | Hnrnpu (113DEL, exon 1) | NA | Impaired (pup calls) | NA | Mild impairment | Impaired gait | Dugger et al., 2023 |
| | | (Δexon 3-6) | NA | NA | NA | NA | Impaired locomotor activity | Lai et al., 2018 |
| ***KMT2E*** | Complex NDD<br>- SFARI: 1S | Kmt2e (8DEL, exon 3) | Impaired | NA | Impaired | NA | Anxiety | Li et al., 2023 |

| | | | | | | | | |
|---|---|---|---|---|---|---|---|---|
| (7q22.3) | - EAGLE: 38.65 | | | | | | | |
| ***TCF20*** (22q13.2) | Developmental delay with variable intellectual disability and behavioural abnormalities - SFARI score: 1 - EAGLE score: 38.0 | Tcf20 (Δexon 2) | Impaired | Impaired | Impaired | Impaired | Normal locomotion activity Anxiety | Feng et al., 2020 |
| ***AUTS2*** (7q11.22) | Syndromic intellectual disability - SFARI: 1 - EAGLE: 35.5 | Auts2 (Δexon 8) | Impaired | Impaired | NA | NA | Impaired sensorimotor gating | Hori et al., 2020 |
| | | Auts2 (En1-Cre, Δexon 8) | NA | Impaired | NA | NA | Impaired | Yamashiro et al., 2020 |
| | | Auts2 (Emx1-Cre, Δexon 8) | Impaired | NA | Impaired | NA | NA | Liu et al., 2023 |
| ***POGZ*** (1q21.3) | Intellectual disability-microcephaly-strabismus-behavioral abnormalities syndrome - SFARI score: 1 - EAGLE score: 35.4 | Pogz (Δexon 13-19) | Normal | NA | Normal | Impaired | Impaired motor coordination Normal locomotion activity | Suliman-Lavie et al., 2020 |
| | | Pogz (Δexons 1 and 6) | Normal | NA | NA | Normal | Normal locomotion activity Anxiolytic | Cunniff et al., 2020 |
| | | $Pogz^{Q1038R/+}$ | Impaired | Impaired (isolated pups) | Impaired | Impaired | Normal locomotion activity Anxiolytic | Matsumura et al., 2020 |
| ***MED13L*** (17q23.2) | Syndromic intellectual disability - SFARI: 1S - EAGLE: 9.3 | Med13l (Δexon 2) | Normal | NA | Normal | Impaired | Impaired motor coordination | Li et al., 2025 |
| ***ARID1B*** (6q25.3) | Coffin-Siris syndrome - SFARI score: 1 - EAGLE score: 34.75 | Arid1b (Δexon 5) | Mild impairment (social novelty) | Impaired (isolated pups) | Impaired | Impaired | Impaired grip strength Anxiety | Jung et al., 2017; Celen et al., 2017 |
| | | Arid1b (Dlx5/6-Cre KO) | Impaired | NA | Impaired | Impaired | Hypoactivity Anxiety | Jung et al., 2017 |
| ***PTCHD1-AS*** (Xp22.11) | NA - SFARI: 2 - EAGLE: 17.6 | *$Ptchd1\text{-}as^{-Ex3/Y}$* (Exon3 KO) | Impaired | Impaired | Impaired | Normal | Normal | Bradley et al., 2026 |
| | | *$Ptchd1\text{-}as^{PolyA/Y}$* (PolyA KI) | Impaired | Impaired | Impaired | Normal | Normal | |
| ***CTNNB1*** (3p22.1) | CTNNB1-related neurodevelopmental disorder and/or vitreoretinopathy - SFARI: 1 - EAGLE: 32.75 | Ctnnb1 (Δexon 2-6) | NA | NA | NA | Impaired | Impaired | Alexander et al., 2024 |
| | | Ctnnb1 (PV-Cre, Δexon 2-6) | Mild impairment (social novelty) | NA | Impaired | Enhanced | Anxiety | Dong et al., 2016 |
| ***NAA15*** (4q31.1) | Syndromic intellectual disability - SFARI: 1S - EAGLE: 31.7 | Naa15 (Δexon 2-6) | Normal | NA | Impaired | NA | Hyperactivity Anxiety | He et al., 2025 |
| ***CREBBP*** (16p13.3) | Rubinstein-Taybi syndrome - SFARI: 1S - EAGLE: 31.35 | $Cbp^{stop523}$ (CaMKIIα-Cre, Δexon 7) | Mild impairment (social novelty) | NA | NA | Impaired | Normal | Lipinski et al., 2022 |

| | | | | | | | | |
|---|---|---|---|---|---|---|---|---|
| | | Cbp (ΔCH1 domain) | Impaired | NA | Impaired | Mild impairment | Hyperactivity Anxiety | Zheng et al., 2016 |
| | | CbpΔ1 (Truncated N-terminal overexpression) | NA | NA | NA | Impaired | Normal | Wood et al., 2005 |
| ***SLC6A1*** (3p25.3) | Complex NDD - SFARI: 1S - EAGLE: 31.15 | Slc6a KO (Gat1 KO) | Impaired (interaction↑) | NA | NA | Impaired | Hyperactivity Impaired motor coordination Impaired sensorimotor gating | Gong et al., 2009; Yu et al., 2013; Chen et al., 2015; Guo et al., 2025 |
| | | Slc6a1[S295L] | NA | NA | NA | Impaired | Impaired motor coordination | |
| | | Slc6a1[A288V] | NA | NA | NA | Mild impairment | Normal | |
| | | Slc6a (Δexon 2-3) | NA | NA | NA | Impaired | Hyperactivity | Gong et al., 2009 |
| ***DEAF1*** (11p15.5) | NA - SFARI: 1S - EAGLE: 30.9 | Deaf1 KO | NA | NA | NA | NA | Anxiety | Luckhart et al., 2016 |
| | | Deaf1 (Δexon 2-5) | NA | NA | NA | Impaired | NA | Silfhout et al., 2014 |
| | | Deaf1 (Nestin-Cre, Δexon 2-5) | NA | NA | NA | Impaired | Mild hypoactivity | |
| ***GRIN2B*** (12p13.1) | Complex NDD - SFARI: 1 - EAGLE: 29.65 | Grin2b[C456Y] (Δexon 6) | Normal | Normal (pup call: impaired) | Mild impairment | Mild impairment | Hypoactivity Anxiolytic | Shin et al., 2020; Kang et al., 2025 |
| ***ASXL3*** (18q12.1) | Syndromic intellectual disability - SFARI: 1S - EAGLE: 28.85 | NA | NA | NA | NA | NA | NA | NA |
| ***SETD5*** (3p25.3) | Syndromic complex NDD - SFARI score: 1 - EAGLE score: 28.05 | Setd5 (Δexon 1) | Mild impairment (social novelty) | NA | NA | Impaired | Hyperactivity Anxiety | Moore et al., 2019 |
| | | Setd5 (Δexon 3-6) | Normal | Impaired (pup calls) | Normal | Mild impairment (extinction) | Normal locomotor activity Normal anxiety | Deliu et al., 2018 |
| ***TRIP12*** (2q36.3) | Complex NDD - SFARI: 1S - EAGLE: 27 | NA | NA | NA | NA | NA | NA | NA |
| ***CHD2*** (15q26.1) | Complex NDD - SFARI: 1S - EAGLE: 25 | Chd2 (β-actin-Cre, Δexon 3) | NA | NA | NA | Impaired | Normal | Kim et al., 2018 |
| | | (Actb-Cre, Δexon 3) | Mild impairment (social novelty) | NA | Normal | Impaired | Normal | Yoon et al., 2026 |
| | | Chd2 (266-267DEL) | Mild impairment | NA | Impaired | NA | Impaired motor coordination | Mavashov et al., 2025 |

| | | | | | | | | |
|---|---|---|---|---|---|---|---|---|
| ***SATB2*** (2q33.1) | SATB2 associated disorder<br>- SFARI: 1S<br>- EAGLE: 24.95 | Satb2 KO<br>(Emx1-Cre) | Mild impairment<br>(interaction ↑)<br>(novelty ↓) | NA | NA | Impaired | Hyperactivity<br>Impaired sensorimotor gating<br>Anxiolytic | Zhang et al., 2019 |
| ***TANC2*** (17q23.2-q23.3) | Intellectual developmental disorder with autistic features and language delay, with or without seizures<br>- SFARI: 1S<br>- EAGLE: 24.95 | Tanc2<br>(Δintron 4) | Normal | Impaired<br>(pup calls) | NA | Impaired | Hyperactivity<br>Anxiolytic | Kim et al., 2021 |
| ***CTTNBP2*** (7q31.31) | NA<br>- SFARI: 2<br>- EAGLE: 26.8 | Cttnbp2<br>(Δexon 3) | Impaired | Impaired<br>(Cttnbp2$^{R533*}$) | Normal | Impaired | Hyperactivity<br>Anxiolytic | Shih et al., 2020 |
| ***NCKAP1*** (2q32.1) | Complex NDD<br>- SFARI: 1<br>- EAGLE: 24.75 | NA | NA | NA | NA | NA | NA | NA |
| ***EP300*** (22q13.2) | Rubinstein-Taybi syndrome due to EP300 haploinsufficiency<br>- SFARI: 1S<br>- EAGLE: 23.6 | Ep300<br>(Δexon 4-6) | NA | NA | NA | Mild impairment<br>(reversal learning) | Normal locomotor activity<br>Anxiety | Viosca et al., 2010 |
| ***NSD1*** (5q35.3) | Sotos syndrome<br>- SFARI: 1S<br>- EAGLE: 23.4 | Nsd1<br>(Δexon 3) | Mild impairment<br>(social novelty) | Impaired<br>(pup calls) | NA | Normal | Normal | Oishi et al., 2020 |
| ***ANKRD11*** (16q24.3) | KBG syndrome<br>- SFARI: 1S<br>- EAGLE: 22.6 | Ankrd1l<br>(ΔC-terminal repression domain) | Impaired | NA | Impaired | NA | Hypoactivity | Gallagher et al., 2015 |
| | | Ankrd11$^{Y347A}$ | Mild impairment<br>(social novelty) | NA | Impaired | Impaired | Normal | Liu et al., 2025 |
| ***MYT1L*** (2p25.3) | Syndromic complex NDD<br>- SFARI: 1<br>- EAGLE: 20.35 | Myt1l<br>(Δexon 6) | Mild impairment<br>(social novelty) | Mild impairment<br>(pup calls) | Normal | Mild impairment | Hyperactivity<br>Impaired motor coordination<br>Anxiolytic | Wohr et al., 2022; Weigel et al., 2023 |
| | | Myt1l<br>(Δexon 7) | Mild impairment<br>(sociability) | Impaired<br>(isolated pups) | Normal | Mild impairment | Hyperactivity<br>Normal locomotor activity<br>Normal sensorimotor gating | Chen et al., 2021 |
| | | Myt1l<br>(Δexon 9) | Mild impairment<br>(sociability) | Normal | Mild impairment | Mild impairment | Hyperactivity<br>Anxiolytic | Kim et al., 2022 |
| ***DYRK1A*** (21q22.13) | Complex NDD<br>- SFARI: 1S<br>- EAGLE: 20.2 | Dyrkla<br>(Δexon 3) | Impaired | Mild impairment<br>(pup calls) | Normal | Mild impairment<br>(reversal learning) | Normal | Raveau et al., 2018 |
| | | Dyrkla<br>(Δintron 4) | Mild impairment<br>(social novelty) | NA | NA | NA | Normal locomotor activity<br>Anxiety | Lin et al., 2026 |
| | | Dyrkla<br>(EIIa-Cre; Δexon 5-6) | Mild impairment<br>(sociability) | NA | NA | Normal | Normal locomotor activity | Shih et al., 2023 |
| ***MARK2*** (11q13.1) | NA<br>- SFARI: 2<br>- EAGLE: 20.05 | Mark2<br>(Δexon 2-17) | Impaired | NA | Impaired | Impaired | Anxiety | Gong et al., 2024 |

| | | | | | | | | |
|---|---|---|---|---|---|---|---|---|
| ***STXBP1***<br>(9q34.11) | Genetic developmental and epileptic encephalopathy<br>- SFARI: 1S<br>- EAGLE: 19.1 | Stxbp1<br>(Δexon 3) | Mild impairment<br>(aggression↑) | NA | NA | Impaired | Normal locomotor activity<br>Anxiety | Miyamoto et al., 2017 |
| | | Stxbp1<br>(Δexon 2-6) | Normal | NA | Mild impairment | Impaired | Hyperactivity<br>Anxiety | Kovacevic et al., 2018 |
| ***CIC***<br>(19q13.2) | NA<br>- SFARI: 1<br>- EAGLE: 18.7 | Cic<br>(forebrain KO) | Normal | NA | NA | Impaired | Hyperactivity<br>Anxiolytic | Lu et al., 2017 |
| | | Cic<br>(hypothalamus and amygdala KO) | Mild impairment<br>(sociability) | NA | NA | Normal | Normal anxiety | |
| ***KMT2A***<br>(11q23.3) | Wiedemann-Steiner syndrome<br>- SFARI: 1S<br>- EAGLE: 18.55 | Mll1<br>(CaMKIIα-Cre; Δexon 3-4) | NA | NA | NA | Impaired | Hyperactivity<br>Anxiety | Jakovcevski et al., 2015; Shen et al., 2016 |
| ***SHANK2***<br>(11q13.3-q13.4) | Complex NDD<br>- SFARI: 1<br>- EAGLE: 18.55 | Shank2<br>(Δexon 6-7) | Impaired | Impaired | Impaired | Impaired | Hyperactivity<br>Anxiety | Won et al., 2012 |
| | | Shank2<br>(Δexon 7) | Mild impairment<br>(resident intruder) | Impaired | Impaired | Normal | Hyperactivity<br>Normal motor coordination<br>Ataxia<br>Anxiety | Schmeisser et al., 2012 |
| ***UBR5***<br>(8q22.3) | NA<br>- SFARI: 2<br>- EAGLE: 18.45 | NA | NA | NA | NA | NA | NA | NA |
| ***CUL3***<br>(2q36.2) | Complex NDD<br>- SFARI: 1<br>- EAGLE: 18.4 | Cul3<br>(Δexon 6) | Impaired | NA | Normal | Mild impairment | Hyperactivity | Amar et al., 2021 |
| | | Cul3<br>(Δexon 4-7 in forebrain) | Impaired | NA | Normal | NA | Hyperactivity<br>Impaired sensorimotor gating | Rapanelli et al., 2021 |
| | | Cul3<br>(Δexon 4-7;<br>GFAP-Cre) | Impaired | NA | Normal | Normal | Normal locomotor activity<br>Anxiety | Dong et al., 2020 |
| | | Cul3<br>(Δexon 4-7;<br>Nex-Cre) | Impaired | NA | NA | NA | Normal locomotor activity<br>Anxiety | |
| ***DIP2A***<br>(21q22.3) | Complex NDD<br>- SFARI: 1<br>- EAGLE: 18.2 | Dip2a KO | Mild impairment<br>(social novelty) | Impaired | Impaired | Normal | Anxiety | Ma et al., 2019; Jun et al., 2023 |
| ***SOX5***<br>(12p12.1) | Lamb-Shaffer syndrome<br>- SFARI: 1S<br>- EAGLE: 17.6 | NA | NA | NA | NA | NA | NA | NA |
| ***GIGYF1***<br>(7q22.1) | Complex NDD<br>- SFARI: 1<br>- EAGLE: 17.25 | Gigyf1<br>(Δexon 1-9;<br>Nestin-Cre) | Impaired | NA | Mild impairment | Impaired | Normal locomotor activity<br>Anxiety | Chen et al., 2022 |
| | | (Δexon 10-24;<br>CMV-Cre) | Mild impairment<br>(social novelty) | NA | Impaired | Normal | Hypoactivity | Ding et al., 2023 |
| | | (Δexon 10-24;<br>NEX-Cre) | Mild impairment<br>(social novelty) | NA | Impaired | Normal | Normal | |

| | | | | | | | | |
|---|---|---|---|---|---|---|---|---|
| | | (Δexon 10-24; GAD2-Cre) | Normal | NA | Impaired | Impaired | Anxiety | |
| ***WDFY3*** (4q21.23) | Syndromic intellectual disability - SFARI: 1 - EAGLE: 17.2 | NA | NA | NA | NA | NA | NA | NA |
| ***TSC1*** (9q34.13) | Tuberous sclerosis - SFARI: 1S - EAGLE: 17.1 | Tsc1 (L7/Pcp2-Cre KO) | Impaired | Impaired (pup calls) | Impaired | Mild impairment (reversal learninig) | Normal locomotor activity Impaired motor coordination Impaired gait | Tsai et al., 2012 |
| ***RFX3*** (9p24.2) | Complex NDD - SFARI: 1 - EAGLE: 15.95 | Rfx3 (Δexon 10) | NA | NA | NA | NA | Hearing loss | Elkon et al., 2015 |
| ***DNMT3A*** (2p23.3) | Tatton-Brown-Rahman overgrowth - SFARI: 1S - EAGLE: 15.9 | Dnmt3a (Δexon 19; CMV-Cre) | Normal | Impaired (isolated pups) | Impaired | Enhanced | Hypoactivity Anxiety | Christian et al., 2020 |
| | | Dnmt3a$^{P900L}$ | Normal | Impaired (pup calls) | NA | Impaired | Normal locomotor activity | Beard et al., 2023 |
| | | Dnmt3a$^{R878H}$ | Mild impairment (aggression↑) | Impaired (pup calls) | NA | Mild impairment | Normal locomotor activity | |
| ***NIPBL*** (5p13.2) | Cornelia de Lange syndrome - SFARI: 1S - EAGLE: 15.8 | Nipbl$^{564}$ (Δintron 1) | NA | NA | Impaired | NA | Seizure | Kawauchi et al., 2009 |
| ***ASTN2*** (9q33.1) | Complex NDD - SFARI: 2 - EAGLE: 15.75 | Astn2 (Δexon 1) | Impaired | Impaired (pup calls) | Impaired | Mild impairment | Hyperactivity Anxiolytic | Hanzel et al., 2024 |
| | | (Δexon 7) | Mild impairment (interaction↑) | NA | Mild impairment | Mild impairment | Hyperactivity Anxiolytic | Ito et al., |
| ***CSDE1*** (1p13.2) | Complex NDD - SFARI: 1S - EAGLE: 15.55 | NA | NA | NA | NA | NA | NA | NA |
| ***WAC*** (10p12.1) | DeSanto-Shinawi syndrome - SFARI: 1S - EAGLE: 15 | NA | NA | NA | NA | NA | NA | NA |
| ***SHANK1*** (19q13.33) | Complex NDD - SFARI: 2 - EAGLE: 14.65 | Shank1 (exon 14-15 deletion) | Mild impairment (*The results are controversial between studies.) | Impaired | Normal | Impaired | Hypoactivity Impaired motor coordination Mild anxiety | Hung et al., 2008 Silverman et al., 2011 Wöhr et al., 2011 |
| | | Shank1 (R882H KI) | Mild impairment (social novelty) | NA | Impaired | Normal | Normal locomotion activity Normal anxiety | Qin et al., 2022 |
| ***AHDC1*** (1p36.11-p35.3) | AHDC1-related intellectual disability-obstructive sleep apnea-mild dysmorphism syndrome - SFARI: 1S - EAGLE: 14.25 | NA | NA | NA | NA | NA | NA | NA |

| | | | | | | | | |
|---|---|---|---|---|---|---|---|---|
| ***ARHGEF9*** (Xq11.1-q11.2) | X-linked complex NDD<br>- SFARI: 1S<br>- EAGLE: 14.2 | Arhgef9 (Δexon 5; hSyn-Cre in mPFC) | Normal | Impaired | Normal | NA | Normal locomotor activity | Papadopoulos et al., 2007; Jung et al., 2025 |
| ***ASH1L*** (1q22) | Syndromic complex NDD<br>- SFARI: 1<br>- EAGLE: 14.15 | Ash1l (Δexon 2) | Mild impairment (social novelty) | Impaired (pup calls) | Impaired | Impaired | Normal locomotor activity<br>Anxiety | Yan et al., 2022 |
| | | Ash1l (Δexon 4; Nestin-Cre) | Impaired | NA | Impaired | Impaired | Hyperactivity | Gao et al., 2021 |
| ***KMT2C*** (7q36.1) | Syndromic intellectual disability<br>- SFARI: 1S<br>- EAGLE: 14.1 | Kmt2c (Δexon 3) | Mild impairment | NA | Impaired | Impaired | Normal locomotor activity | Brauer et al., 2023 |
| | | (Δexon 14) | Mild impairment | NA | Impaired | Impaired | Normal locomotor activity | Ma et al., 2025 |
| | | (Δexon 43) | Mild impairment | NA | NA | Impaired | Impaired sensorimotor gating | Nakamura et al., 2024 |
| ***KMT5B*** (11q13.2) | Complex NDD<br>- SFARI: 1<br>- EAGLE: 14.05 | NA | NA | NA | NA | NA | NA | NA |
| ***MSL3*** (Xp22.2) | Basilicata-Akhtar syndrome<br>- SFARI: 1S<br>- EAGLE: 13.9 | NA | NA | NA | NA | NA | NA | NA |
| ***ARX*** (Xp21.3) | X-linked complex NDD<br>- SFARI: 1S<br>- EAGLE: 13.8 | $Arx^{dup24/0}$/ $Arx^{(GCG)10+7}$ (Δexon2) | Normal/ Impaired | NA | Impaired | Impaired | Hyperactivity (Normal/Anxiolytic) | Price et al., 2009; Dubos et al., 2018 |
| ***CNTN6*** (3p26.3) | Complex NDD<br>- SFARI: 2<br>- EAGLE: 13.8 | Cntn6 (Δexon2) | Impaired | NA | Normal | NA | NA | Zhang et al., 2023 |
| ***KDM6B*** (17p13.1) | Syndromic intellectual disability<br>- SFARI: 1<br>- EAGLE: 13.75 | Kdm6b (Δexon6) | Impaired | NA | Impaired | Impaired | Hyperactivity<br>Anxiolytic | Brauer et al., 2024 |
| | | Kdm6b KO | Impaired | NA | Normal | Impaired | Hyperactivity<br>Anxiolytic<br>Impulsive behavior | Gao et al., 2022 |
| ***DSCAM*** (21q22.2) | Autism spectrum disorder<br>- SFARI: 1<br>- EAGLE: 13.5 | Dscam (Δexon1; Nestin-Cre) | Impaired | NA | Normal | NA | Normal locomotor activity | Lim et al., 2021 |
| | | (Δexon2-33; GFAP-Cre) | Mild impairment (social novelty) | NA | Impaired | Mild impairment | Mild hyperactivity<br>Anxiety | Chen et al., 2022 |
| | | (Δexon2-33; NEX-Cre) | Mild impairment (social novelty) | NA | Impaired | Mild impairment | Normal locomotor activity<br>Mild anxiety | |
| | | (Δexon17) | NA | NA | NA | Impaired motor learning | Impaired motor coordination | Xu et al., 2011 |
| | | (Δexon19) | NA | NA | NA | Impaired | Hyperactivity<br>Anxiety<br>Impaired motor coordination | Neff et al., 2024 |

| | | | | | | | | |
|---|---|---|---|---|---|---|---|---|
| ***EHMT1*** (9q34.3) | Kleefstra syndrome - SFARI: 1S - EAGLE: 13.5 | Ehmt1 (Δexon25-26) | Impaired | NA | Impaired | Impaired | Hypoactivity Impaired sensorimotor gating Anxiety | Balemans et al., 2010/2013; Benevento et al., 2017 |
| ***TCF4*** (18q21.2) | Pitt-Hopkins syndrome - SFARI: 1S - EAGLE: 13.5 | Tcf4 (Δexon4) | NA | NA | NA | Impaired | Normal locomotor activity | Badowska et al., 2020 |
| | | Tcf4 (ΔC-terminus DNA-binding domain) | Impaired | Impaired (pup calls) | Impaired | Impaired | Hyperactivity Impaired sensorimotor gating Impaired motor coordination | Kennedy et al., 2016; Cleary et al., 2021 |
| ***DYNC1H1*** (14q32.31) | Neuronopathy, distal hereditary motor - SFARI: 1 - EAGLE: 13.45 | Dync1h1$^{c9052C>T}$ (Δexon46) | NA | NA | NA | Normal | Impaired sensorimotor gating Impaired motor coordination | Ramos et al., 2024 |
| ***TLK2*** (17q23.2) | Complex NDD - SFARI: 1S - EAGLE: 13.5 | NA | NA | NA | NA | NA | NA | NA |
| ***BRAF*** (7q34) | Cardiofaciocutaneous syndrome - SFARI: 1S - EAGLE: 13.05 | Braf (Δexon12; CaMKII-Cre) | NA | NA | NA | Impaired | Normal locomotor activity | Chen et al., 2006 |
| | | Braf$^{Q241R}$ (CaMKII-Cre) | NA | NA | NA | Impaired | Hypoactivity | Moriya et al., 2025 |
| | | Braf$^{K499E}$ (Nestin-Cre) | Normal | NA | NA | Impaired | Normal locomotor activity | Kang et al., 2025 |
| | | Braf$^{K499E}$ (CaMKII-Cre/ vGAT-Cre) | Normal | NA | NA | Normal/Normal | Normal locomotor activity | |
| ***CACNA1D*** (3p21.1) | Complex NDD - SFARI: 2 - EAGLE: 12.7 | Cacna1d$^{A771G}$ | Impaired | NA | Impaired | NA | Hyperactivity Mild anxiety | Ortner et al., 2023 |
| | | Cacna1d$^{Ile772Met}$ (Δexon16) | Normal | NA | Impaired | Normal | Hyperactivity Impaired motor coordination | Stolting et al., 2023 |
| ***RAI1*** (17p11.2) | Smith-Magenis syndrome - SFARI: 1S - EAGLE: 12.25 | Rai1 (Δexon2) | Mild impairment | NA | Impaired | Impaired | Normal locomotor activity | Bi et al., 2007; Rao et al., 2017 |
| | | (Δexon3; Nestin-Cre) | Normal | NA | NA | Impaired | Hypoactivity | Huang et al., 2016 |
| ***GRIA2*** (4q32.1) | NDD with language impairment and behavioral abnormalities - SFARI: 1 - EAGLE: 12 | (Δexon11; Drd1-Cre/ Drd2-Cre) | Mild impairment (sociability) | NA | NA | Impaired | Normal/ Hypoactivity | Shou et al., 2019 |
| ***NLGN4X*** (Xp22.32-p22.31) | X-linked complex NDD - SFARI: 1 - EAGLE: 12 | Nlgn4X KO (Δintron1) | Impaired | Impaired | Impaired | Normal | Normal | Jamain et al., 2008; El-Kordi et al., 2013; Ju et al., 2014 |

### Supplementary Figures

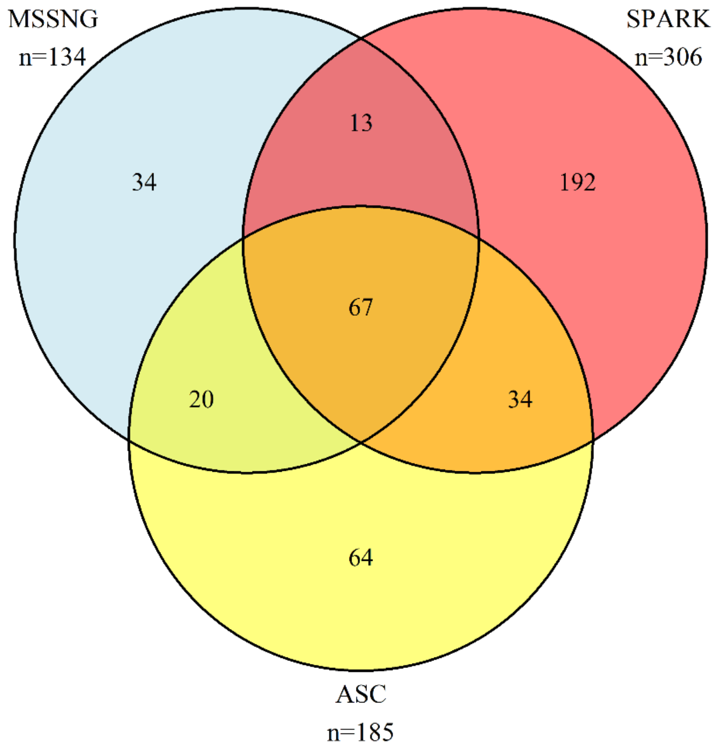


**Supplementary Figure 1 | Overlap of ASD gene lists from large scale genome sequencing resources.** Limited concordance (67/424; 15.8%) among ASD associated genes reported by MSSNG (light blue), SPARK (red) and ASC (yellow).

**a)**

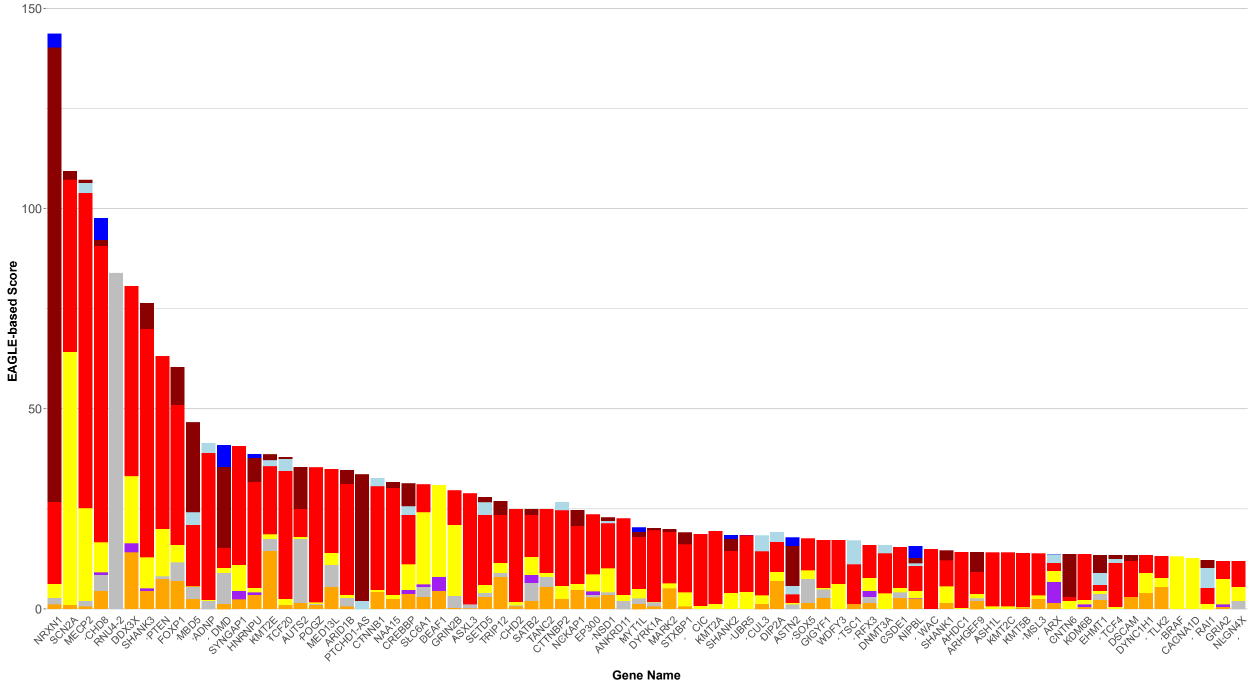
EAGLE-based Score
0
50
100
150
Gene Name
NRXN1
SCN2A
MECP2
CHD8
RNU4-2
DDX3X
SHANK3
PTEN
FOXP1
MBD5
ADNP
DMD
SYNGAP1
HNRNPU
KMT2E
TCF20
AUTS2
POGZ
MED13L
ARID1B
PTCHD1-AS
CTNNB1
NAA15
CREBBP
SLC6A1
DEAF1
GRIN2B
ASXL3
SETD5
TRIP12
CHD2
SATB2
TANC2
CTTNBP2
NCKAP1
EP300
NSD1
ANKRD11
MYT1L
DYRK1A
MARK2
STXBP1
CIC
KMT2A
SHANK2
UBR5
CUL3
DIP2A
ASTN2
SOX5
GIGYF1
WDFY3
TSC1
RFX3
DNMT3A
CSDE1
NIPBL
WAC
SHANK1
AHDC1
ARHGEF9
ASH1L
KMT2C
KMT5B
MSL3
ARX
CNTN6
KDM6B
EHMT1
TCF4
DSCAM
DYNC1H1
TLK2
BRAF
CACNA1D
RAI1
GRIA2
NLGN4X

**b)**

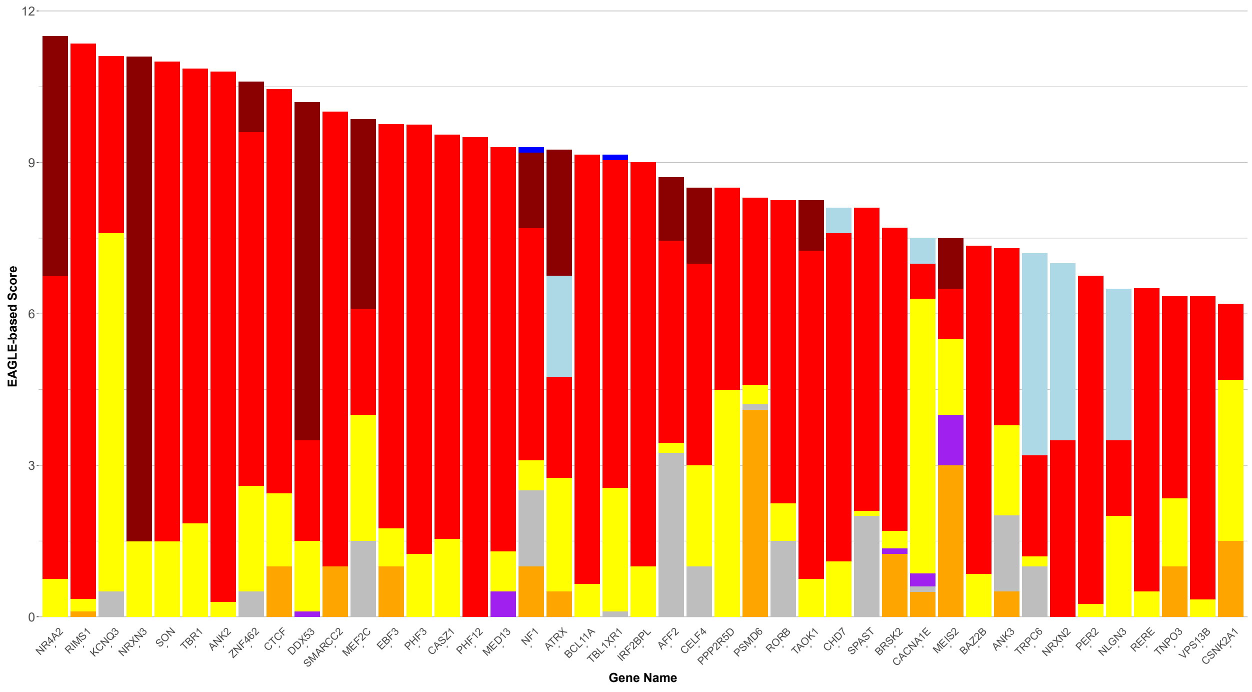
EAGLE-based Score
0
3
6
9
12
Gene Name
NR4A2
RIMS1
KCNQ3
NRXN3
SON
TBR1
ANK2
ZNF462
CTCF
DDX53
SMARCC2
MEF2C
EBF3
PHF3
CASZ1
PHF12
MED13
NF1
ATRX
BCL11A
TBL1XR1
IRF2BPL
AFF2
CELF4
PPP2R5D
PSMD6
RORB
TAOK1
CHD7
SPAST
BRSK2
CACNA1E
MEIS2
BAZ2B
ANK3
TRPC6
NRXN2
PER2
NLGN3
RERE
TNPO3
VPS13B
CSNK2A1

**c)**

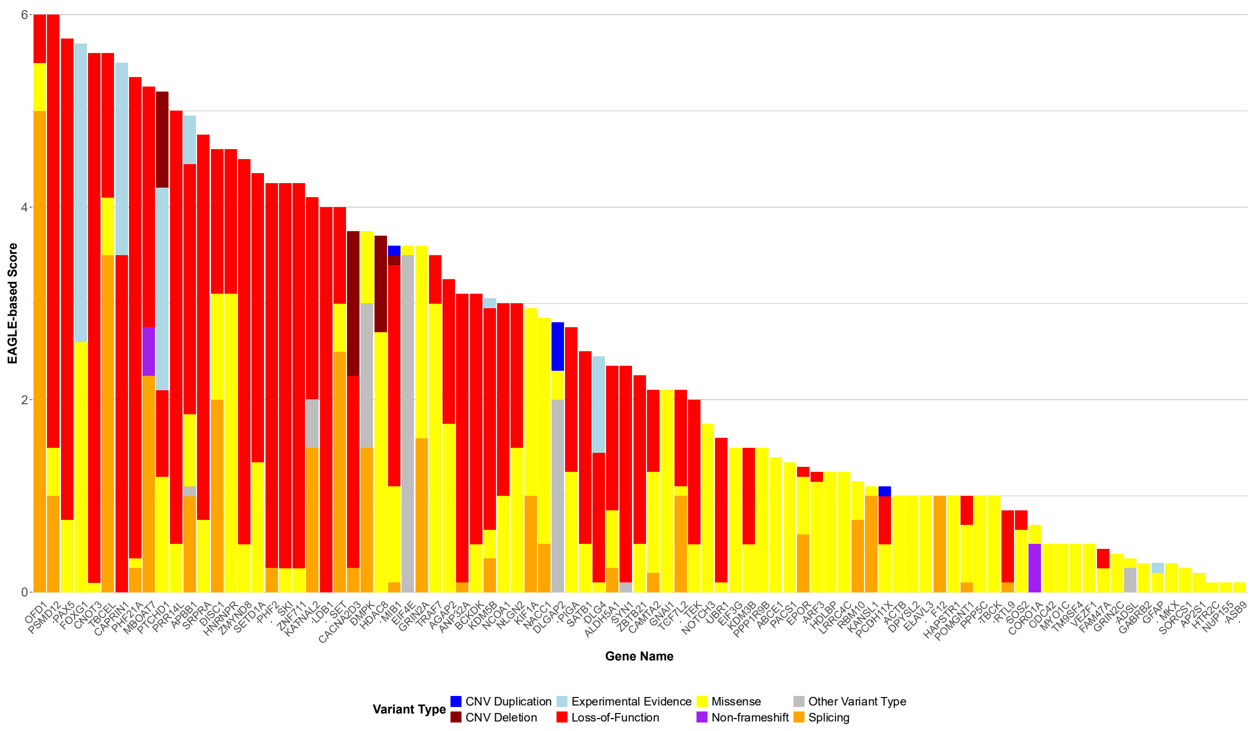


**Supplementary Figure 2 | Detailed characterization of the allelic variation underlying the evidence supporting the association of EAGLE genes with ASD.** The contribution from different variant classes to the EAGLE score was determined for **(a)** EAGLE definitive; **(b)** moderate and **(c)** limited genes.

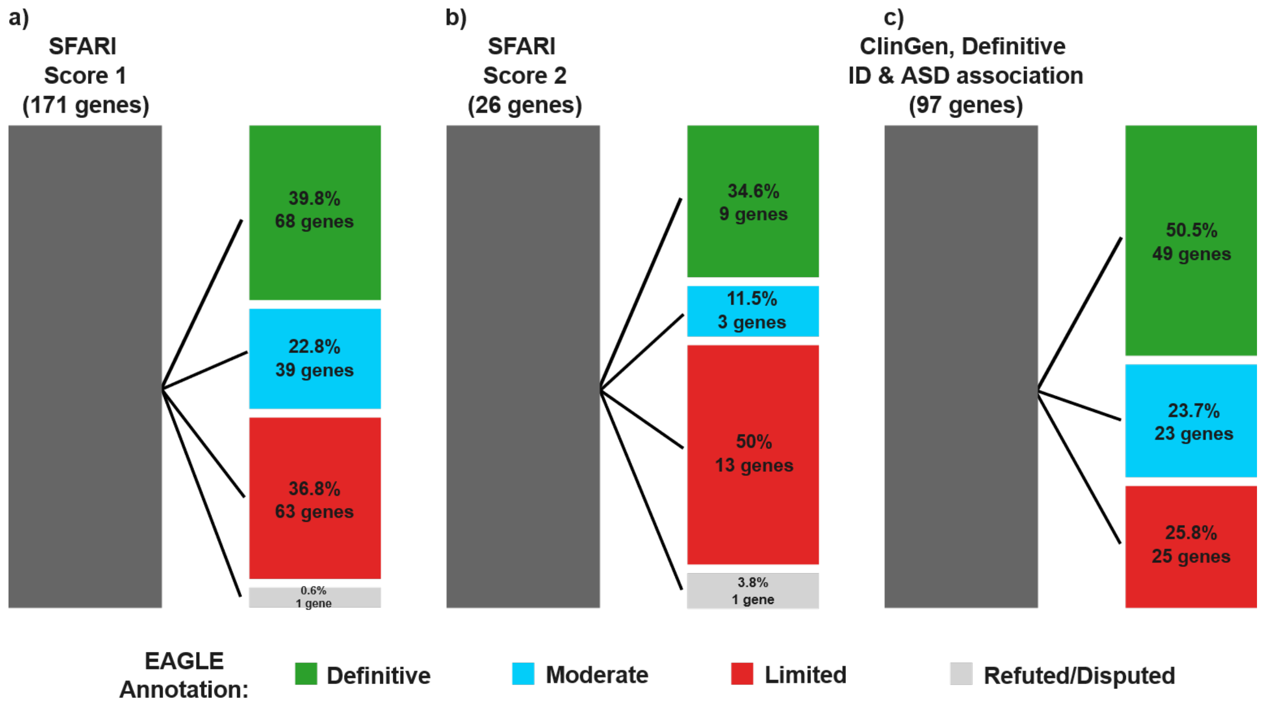


**Supplementary Figure 3 | Discrepancies in the level of gene-ASD associations reported by SFARI-Gene, ClinGen and EAGLE.** Bar plots represent the overlapping genes curated by SFARI-Gene, ClinGen and EAGLE. Distribution of **a)** 171 SFARI Score 1 genes (High confidence), **b)** 26 SFARI Category 2 genes (moderate level of ASD association) and **c)** 97 definitive genes curated by the ClinGen's ID/ASD GCEP based on the EAGLE defined classifications.

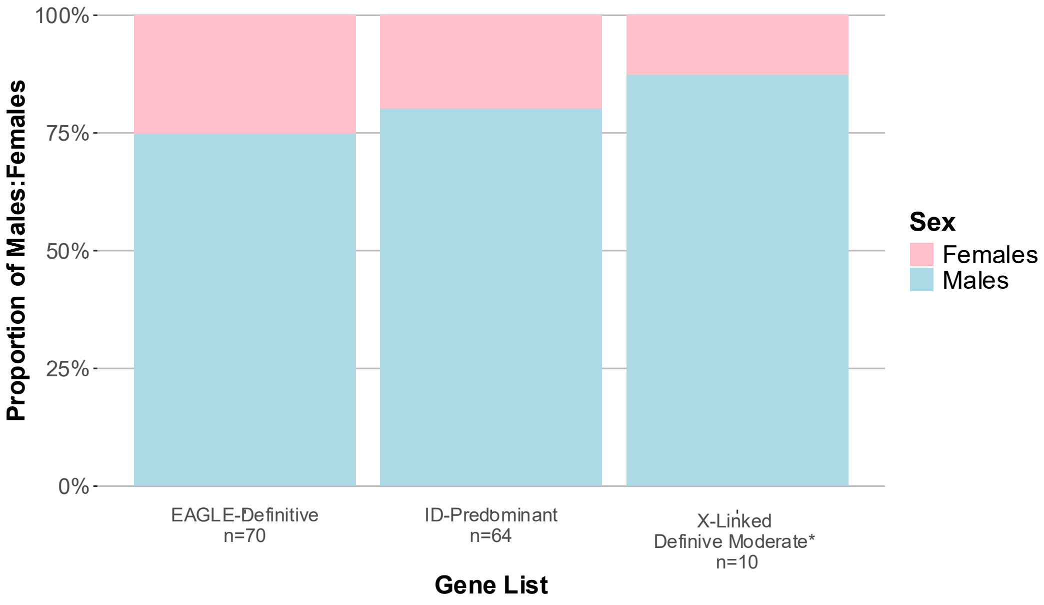


**Supplementary Figure 4 | Proportion of male and female cases included in the case-level evidence assessment process.** Cases stratified by sex included in the curation of EAGLE-definitive, ID-predominant and X-linked, definitive and moderate gene lists. For the first two gene lists we only included autosomal genes. For the X-linked genes we excluded *MECP2* and *DDX3X* since these genes predominantly affect females.

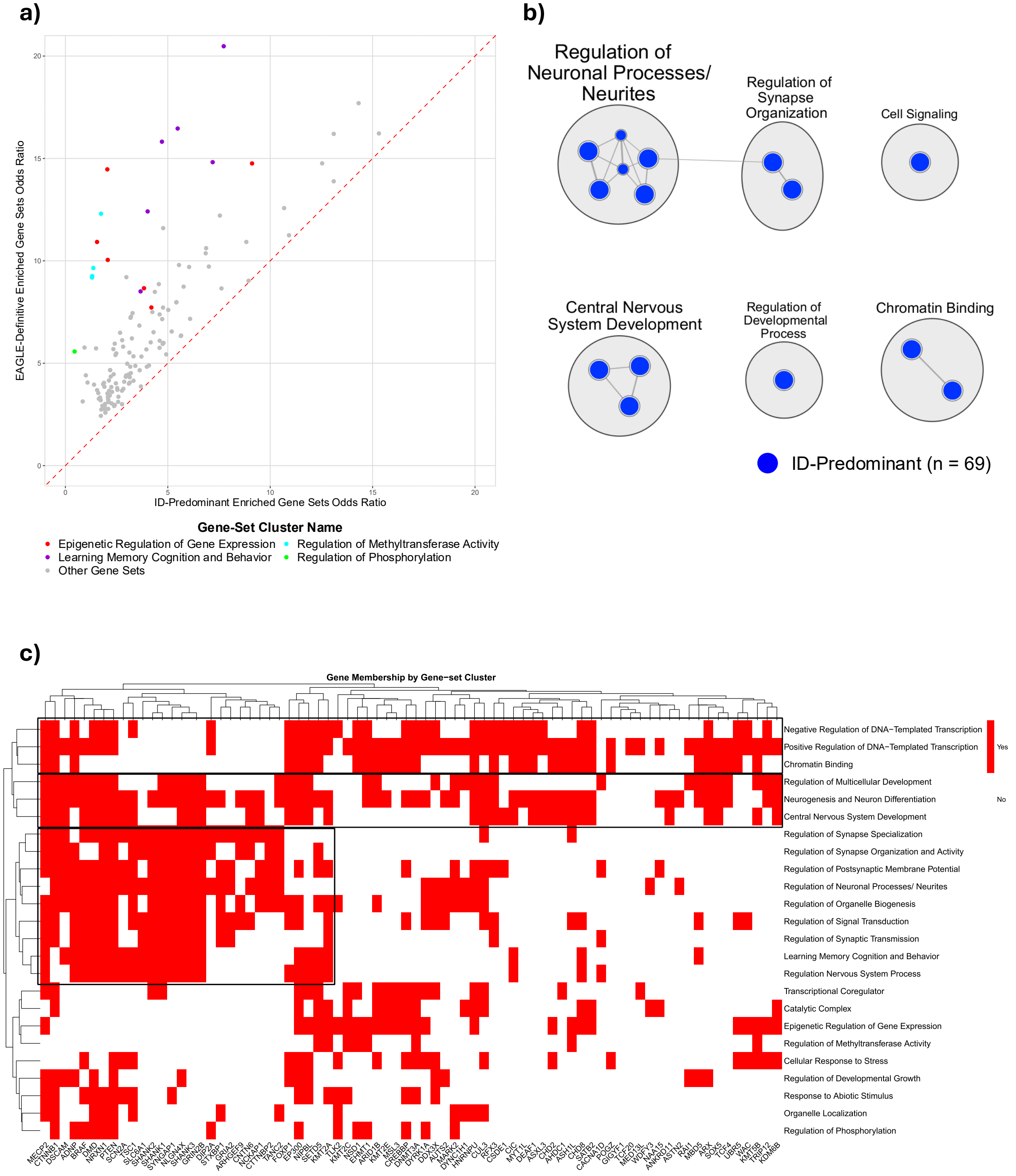


**Supplementary Figure 5 | Differential enrichment of gene sets among EAGLE-Definitive and ID-predominant gene lists. a)** Scatterplot of odds ratios calculated for the ID-predominant (X-axis) and EAGLE-Definitive genes (Y-axis). Odds ratios were calculated for the 141 significant gene sets identified in the enrichment analysis. **b)** Gene sets enriched among the ID-predominant genes. **c)** Detailed mapping of EAGLE-definitive gene to the 24 enriched gene-set clusters.

### Brain Regions Reference

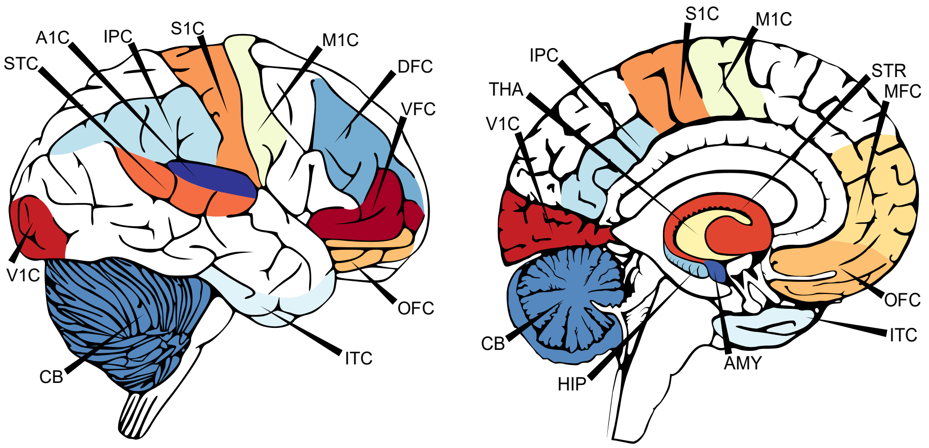


#### Lateral View

#### Sagittal View

- **A1C:** Primary Auditory Cortex
- **AMY:** Amygdala
- **CB:** Cerebellum
- **DFC:** Dorsolateral Prefrontal Cortex
- **HIP:** Hippocampus
- **IPC:** Inferior Parietal Cortex
- **ITC:** Inferior Temporal Cortex
- **M1C:** Primary Motor Cortex
- **THA:** Thalamus
- **MFC:** Medial Prefrontal Cortex
- **OFC:** Orbital Frontal Cortex
- **S1C:** Primary Somatosensory Cortex
- **STC:** Posterior (Caudal) Superior Temporal Cortex
- **STR:** striatum
- **V1C:** Primary Visual Cortex
- **VFC:** Ventrolateral Prefrontal Cortex

**Supplementary Figure 6 | Reference map of brain structures.** Top: Lateral and midsagittal views of the 16 brain structures with available gene expression data after quality control and compatible with the cerebroViz package. Bottom: Brain structures names and acronyms.
